# Physical activity, fatty acids, and MASLD risk: Behavioural and metabolic factors jointly shaping liver health in populations

**DOI:** 10.64898/2026.06.05.26354982

**Authors:** Fangjun Chen, Ruijia You, Yi Liu, Yuheng Yin, Anbin Liu, Lanzhi Deng, Biao Xie, Jie Fan, Wei Wang

**Author notes:** Contributing authors. These two authors contributed equally to this work (Fangjun Chen, Ruijia You).

## Abstract

**Background and Aims:** MASLD has become the most prevalent chronic liver disease globally. Although MVPA and plasma fatty acids have been individually studied in relation to metabolic health, their independent and combined associations with MASLD incidence remain unclear. We aimed to investigate these associations.

**Methods and Results:** This study included 51,717 UK Biobank participants free of liver disease at baseline, with MVPA measured using wrist-worn accelerometers and plasma fatty acids quantified via NMR. Multivariable-adjusted Cox models and restricted cubic splines were used. Over a median follow-up of 7.8 years, 472 incident cases were identified. In fully adjusted models, meeting recommended MVPA levels together with higher n-6 PUFA concentrations was associated with a 71% lower risk (HR 0.29, 95% CI 0.18–0.45). The MVPA-MASLD association was nonlinear, with risk reduction plateauing at approximately 189 minutes per week. Higher n-6 PUFA was associated with reduced risk, whereas n-3 PUFA showed no significant association.

**Conclusions:** These findings suggest that behavioral and metabolic factors may jointly influence MASLD risk. Further studies in diverse populations are needed to confirm these associations.

## 1 Introduction

Metabolic dysfunction-associated steatotic liver disease (MASLD) has become the most common chronic liver disease globally, affecting more than one-third of the adult population worldwide(Chan et al. 2023).MASLD is not only a leading cause of chronic liver disease but also a significant contributor to liver-related morbidity and mortality(Miao et al. 2024).Furthermore, mortality among affected individuals is driven predominantly by cardiovascular disease and cancer, situating MASLD within broader cardiometabolic risk processes rather than as an isolated hepatic condition(Machado 2023).Its increasing prevalence therefore raises questions not only of clinical management but also of population-level risk governance and long-term health system sustainability.

Research on MASLD has largely proceeded within a biomedical paradigm emphasising obesity, insulin resistance, and lipid dysregulation(Truong and Lee 2025; Bansal and Bansal 2024; Lee et al. 2025). While these mechanisms remain central, they capture only part of the relevant risk architecture. Health behaviours and metabolic states are socially patterned(Mudd et al. 2024; Wang et al. 2022).Physical activity opportunities, dietary exposures, and access to preventive resources are shaped by income, occupational structures, neighbourhood environments, and commercial food systems(Jones et al. 2024; Summerhayes et al. 2024).Recognising this socioeconomic patterning provides essential context for interpreting behavioural and metabolic determinants without reducing them to purely individual-level phenomena.Within this broader framework, moderate-to-vigorous physical activity (MVPA) and circulating fatty acid profiles represent two modifiable and potentially interrelated components of MASLD risk. Guidelines issued by the World Health Organization and the American Heart Association recommend that adults accumulate at least 150 minutes of MVPA per week, emphasising the importance of both duration and intensity(Bull et al. 2020; Arnett et al. 2019).Yet attainment of such recommendations varies systematically across socioeconomic contexts. Exercise can improve fatty liver disease through various mechanisms, including increasing fatty acid oxidation and reducing fatty acid synthesis in the liver(Van Der Windt et al. 2018; Sheka et al. 2020).

Evidence regarding associations between specific fatty acid classes and MASLD remains heterogeneous. Some studies suggest that increasing dietary intake of n-6 and n-3 polyunsaturated fatty acids (PUFA) is negatively correlated with MASLD risk(Cui et al. 2021).whereas plasma-based analyses indicate that increased MASLD risk is associated with reduced circulating n-6 PUFA, but not necessarily n-3 PUFA(Frankovic et al. 2024).Such inconsistencies likely reflect, at least in part, limitations inherent in dietary recall instruments, which introduce systematic error in exposure characterisation. Plasma fatty acids, as objective biomarkers quantified through nuclear magnetic resonance (NMR) spectroscopy, can more directly reflect metabolic status and provide a more reliable basis for inference(Masoodi et al. 2021).Two methodological considerations are central to advancing this field. First, the predominant reliance on self-reported physical activity introduces differential misclassification and constrains precise modelling of dose–response relationships(Qiu et al. 2017; Li et al. 2023; Füzéki et al. 2017).The WHO Physical Activity Guidelines Group has explicitly recommended using wearable devices to assess the relationship between physical activity and disease risk(DiPietro et al. 2020).Wearable devices enable objective measurement and long-term continuous monitoring, making them optimal tools for quantifying and tracking activity levels(Li et al. 2025).A recent study reported a non-linear dose-response relationship between accelerometer-measured MVPA and MAFLD(Liu et al. 2024).Second, dietary recall methods introduce analogous measurement error in fatty acid assessment. NMR-based quantification of plasma fatty acids, as implemented in the UK Biobank(UKB), provides objective biomarkers that integrate both dietary intake and endogenous metabolism(Julkunen et al. 2023).

Limited research has examined the integrated analysis of physical activity and fatty acid profiles, despite their behavioural and metabolic interconnections. Drawing on UKB data with device-based physical activity and NMR-based biomarker profiling, this study investigates the independent and joint associations of accelerometer-measured MVPA and plasma fatty acid composition with incident MASLD. By incorporating sociodemographic factors such as the Townsend deprivation index, this analysis aims to enhance methodological rigour in chronic disease epidemiology and inform prevention strategies that account for both behavioural and structural determinants.

## 2 Methods

### 2.1 Study design and participants

The UK Biobank (UKB) is a prospective cohort study that recruited 500,000 participants aged 40–69 years from 22 assessment centres across England, Scotland, and Wales between 2006 and 2010. Baseline data were collected via touchscreen questionnaires, interviews, physical measurements, and biological samples. The study was approved by the North West Multi-Centre Research Ethics Committee (21/NW/0157), and all participants provided written informed consent.

Between February 2013 and December 2015, 103,620 participants wore an accelerometer (Axivity, UK) on their dominant wrist for one week. This study included participants with complete plasma fatty acid data (over 270,000). After data quality control and exclusion of participants with MASLD or severe liver diseases at or before accelerometer measurement, 51,717 participants were included (see Fig. 1). The analysis was conducted under UKB application number 160,957.

**Fig. 1.**
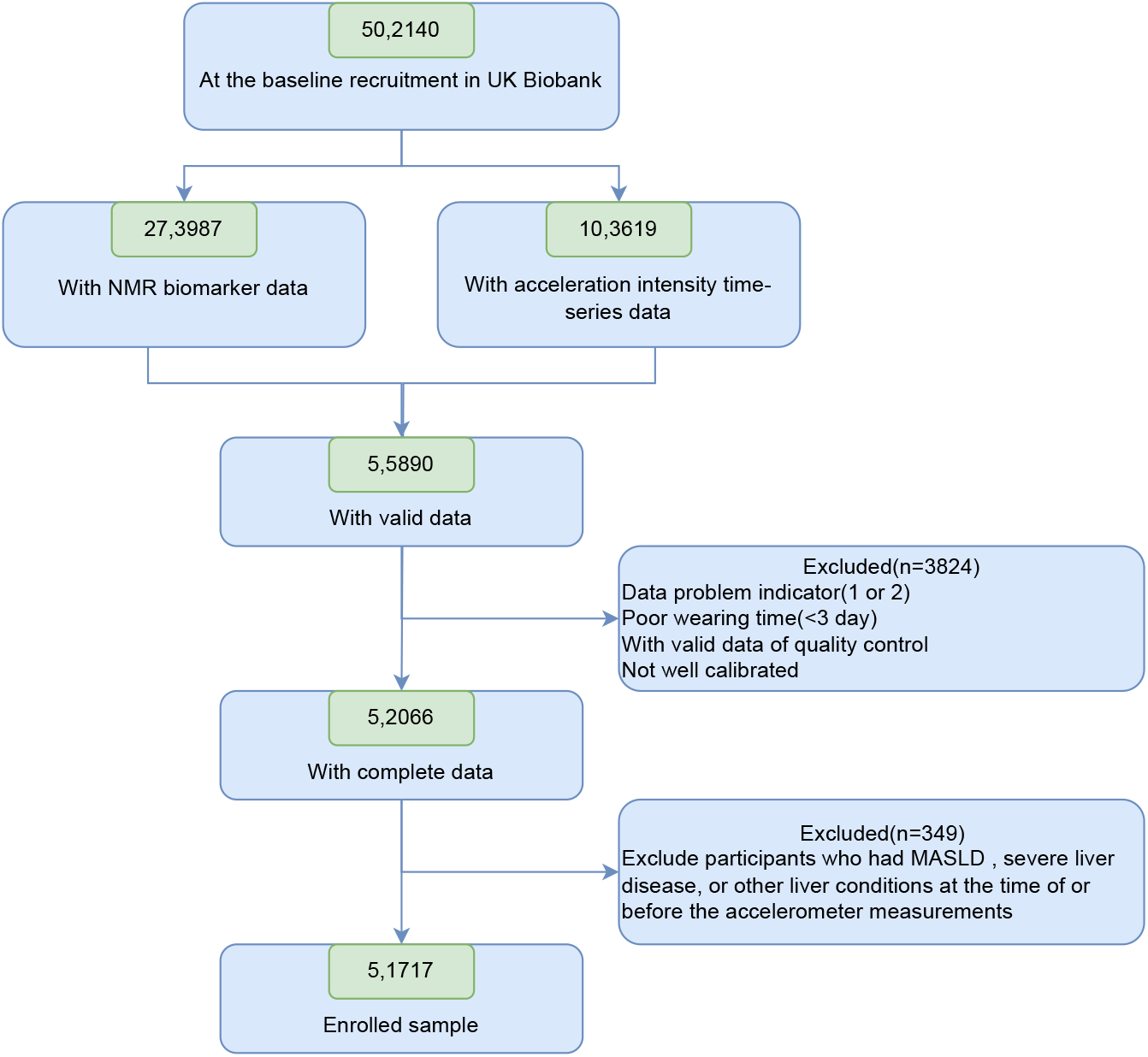
The flowchart of participant enrollment.NMR, nuclear magnetic resonance; MASLD, metabolic dysfunction-associated steatotic liver disease.

### 2.2 Exposures

Exposures included accelerometer-measured MVPA and specific plasma fatty acids. Raw accelerometer data were processed by the UKB working group (Doherty et al. 2017). MVPA was defined using a refined method focusing on brief episodes: at least 80% of 5-second epochs per minute with an average vector magnitude ≥ 100 mg (Li et al. 2025), replacing prior approaches using 5-minute windows (Khurshid et al. 2021; Feng et al. 2023). Accelerometer data validity was confirmed against the doubly labelled water method (White et al. 2019).

Plasma fatty acids (n-3 PUFA, n-6 PUFA, MUFA, SFA) were measured using a high-throughput NMR platform and expressed as percentages of total fatty acids (% TFA). Samples were collected between 2007 and 2010 and stored at − 80^°^C. Metabolomics analysis was conducted in two phases (2019–2022); the first-phase dataset (118,461 samples) was used (Julkunen et al. 2023).

Accelerometer data were excluded for participants with < 3 days (72 hours) of wear time or missing data across the 24-hour cycle, based on UKB working group guidelines (Doherty et al. 2017).

### 2.3 Covariates

Age, sex, ethnicity (White/non-White), Townsend index, assessment centre (England/Scotland/Wales), education (university/other/none), sleep duration, smoking (never/former/current), alcohol (never/< 3 times/week/ ≥ 3 times/week), BMI, season (spring/-summer/autumn/winter), hypertension, platelet count, ALT, AST, cholesterol, triglycerides.

### 2.4 Study outcome

MASLD (K76.0) and MASH (K75.8) were identified from hospital admissions and death registries, with data sources varying by region (Scotland: Scottish Morbidity Records and NHS Central Register; England/Wales: Health Episode Statistics and NHS Information Centre). At analysis (March 1, 2025), admission data were available until October 31, 2022 (England), August 31, 2022 (Scotland), and May 31, 2022 (Wales); death data until November 12, 2021. Follow-up began at accelerometer wear and ended at outcome, death, or centre-specific review date, whichever came first.

### 2.5 Statistical analysis

Continuous variables were presented as median (IQR) and categorical variables as frequency (%). Missing data (all < 5%) were imputed by chained equations. Group differences were assessed by Kruskal-Wallis/rank sum test (continuous) or chisquare/Fisher’s exact test (categorical).

MVPA was categorised by quartiles and the guideline threshold ( ≥ 150 min/week) (Bull et al. 2020). Plasma fatty acids were expressed as % of total fatty acids and divided into quartiles.Multivariable Cox regression was used to estimate hazard ratios (HRs) and 95% CIs for MASLD. Three stepwise-adjusted models were fitted: Model 1 adjusted for age, sex, ethnicity; Model 2 further adjusted for Townsend index, centre, education, sleep, smoking, alcohol, BMI, season; Model 3 additionally adjusted for hypertension, platelets, ALT, AST, cholesterol, triglycerides. Incidence rate was per 1000 person-years. Proportional hazards were verified (Schoenfeld residuals, no violation).Kaplan-Meier curves with log-rank tests compared cumulative incidence. Nonlinear dose–response was assessed using restricted cubic splines (RCS) in Cox models; where nonlinear, a two-segment Cox model was fitted around the inflection point identified by stepwise quantile search.Joint effects were examined by dividing the sample into eight groups based on MVPA recommendation status and fatty acid quartiles, using Cox models with the *Not Recommended* MVPA group combined with n-6 PUFA Q1, MUFA Q4, or SFA Q4 as the reference. Subgroup and interaction analyses were conducted by gender, age, ethnicity, centre, education, smoking, alcohol, BMI, season, and hypertension. Sensitivity analyses were performed.

All analyses used R (version 4.4.2). Two-tailed *P* < 0.05 was considered significant.

## 3 Results

### 3.1 Baseline characteristics

Table 1 presents the baseline characteristics of participants grouped by MVPA quartiles. The study included a total of 51,717 participants, with a median age of 57 years (IQR: 50–62), of which 28,882 (55.8%) were female, and 97.2% were White. During a median follow-up period of 7.8 years (IQR: 7.4–8.3), a total of 472 cases of MASLD occurred. The median MVPA measured by accelerometer was 196 minutes/week (IQR: 108–320), which is higher than the 150 minutes/week recommended by the World Health Organization. Compared to participants with the lowest MVPA levels, those with higher MVPA levels were generally younger, more educated, had lower body mass index (BMI) and waist circumference, and were less likely to smoke or have hypertension.

**Table 1.**
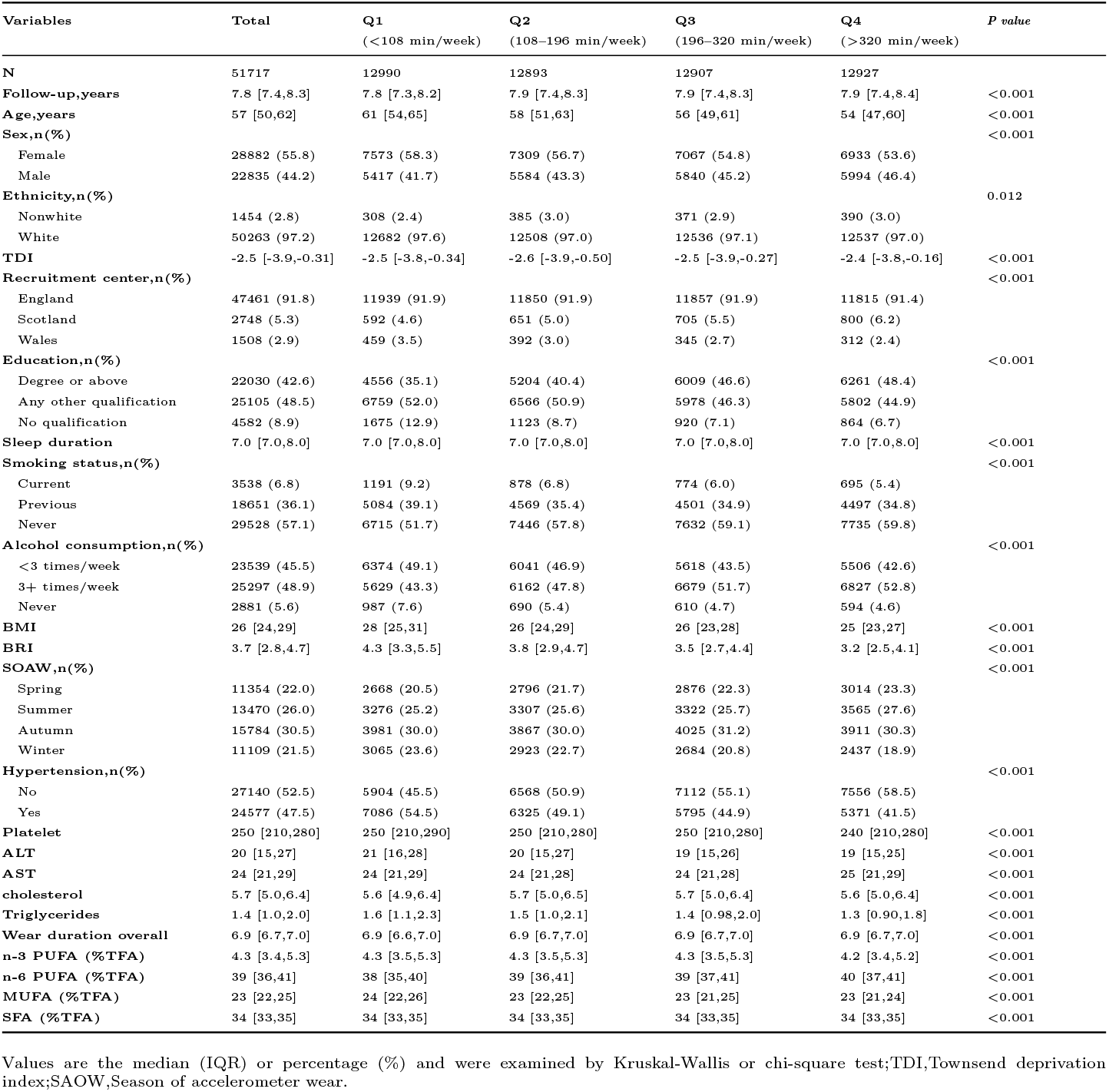
Baseline characteristics by quartiles of MVPA volume groups (n = 51,717).

| Variables | Total | Q1<br>(<108 min/week) | Q2<br>(108–196 min/week) | Q3<br>(196–320 min/week) | Q4<br>(>320 min/week) | *P value* |
| --- | --- | --- | --- | --- | --- | --- |
| **N** | 51717 | 12990 | 12893 | 12907 | 12927 |  |
| **Follow-up,years** | 7.8 [7.4,8.3] | 7.8 [7.3,8.2] | 7.9 [7.4,8.3] | 7.9 [7.4,8.3] | 7.9 [7.4,8.4] | <0.001 |
| **Age,years** | 57 [50,62] | 61 [54,65] | 58 [51,63] | 56 [49,61] | 54 [47,60] | <0.001 |
| **Sex,n(%)** |  |  |  |  |  | <0.001 |
| Female | 28882 (55.8) | 7573 (58.3) | 7309 (56.7) | 7067 (54.8) | 6933 (53.6) |  |
| Male | 22835 (44.2) | 5417 (41.7) | 5584 (43.3) | 5840 (45.2) | 5994 (46.4) |  |
| **Ethnicity,n(%)** |  |  |  |  |  | 0.012 |
| Nonwhite | 1454 (2.8) | 308 (2.4) | 385 (3.0) | 371 (2.9) | 390 (3.0) |  |
| White | 50263 (97.2) | 12682 (97.6) | 12508 (97.0) | 12536 (97.1) | 12537 (97.0) |  |
| **TDI** | -2.5 [-3.9,-0.31] | -2.5 [-3.8,-0.34] | -2.6 [-3.9,-0.50] | -2.5 [-3.9,-0.27] | -2.4 [-3.8,-0.16] | <0.001 |
| **Recruitment center,n(%)** |  |  |  |  |  | <0.001 |
| England | 47461 (91.8) | 11939 (91.9) | 11850 (91.9) | 11857 (91.9) | 11815 (91.4) |  |
| Scotland | 2748 (5.3) | 592 (4.6) | 651 (5.0) | 705 (5.5) | 800 (6.2) |  |
| Wales | 1508 (2.9) | 459 (3.5) | 392 (3.0) | 345 (2.7) | 312 (2.4) |  |
| **Education,n(%)** |  |  |  |  |  | <0.001 |
| Degree or above | 22030 (42.6) | 4556 (35.1) | 5204 (40.4) | 6009 (46.6) | 6261 (48.4) |  |
| Any other qualification | 25105 (48.5) | 6759 (52.0) | 6566 (50.9) | 5978 (46.3) | 5802 (44.9) |  |
| No qualification | 4582 (8.9) | 1675 (12.9) | 1123 (8.7) | 920 (7.1) | 864 (6.7) |  |
| **Sleep duration** | 7.0 [7.0,8.0] | 7.0 [7.0,8.0] | 7.0 [7.0,8.0] | 7.0 [7.0,8.0] | 7.0 [7.0,8.0] | <0.001 |
| **Smoking status,n(%)** |  |  |  |  |  | <0.001 |
| Current | 3538 (6.8) | 1191 (9.2) | 878 (6.8) | 774 (6.0) | 695 (5.4) |  |
| Previous | 18651 (36.1) | 5084 (39.1) | 4569 (35.4) | 4501 (34.9) | 4497 (34.8) |  |
| Never | 29528 (57.1) | 6715 (51.7) | 7446 (57.8) | 7632 (59.1) | 7735 (59.8) |  |
| **Alcohol consumption,n(%)** |  |  |  |  |  | <0.001 |
| <3 times/week | 23539 (45.5) | 6374 (49.1) | 6041 (46.9) | 5618 (43.5) | 5506 (42.6) |  |
| 3+ times/week | 25297 (48.9) | 5629 (43.3) | 6162 (47.8) | 6679 (51.7) | 6827 (52.8) |  |
| Never | 2881 (5.6) | 987 (7.6) | 690 (5.4) | 610 (4.7) | 594 (4.6) |  |
| **BMI** | 26 [24,29] | 28 [25,31] | 26 [24,29] | 26 [23,28] | 25 [23,27] | <0.001 |
| **BRI** | 3.7 [2.8,4.7] | 4.3 [3.3,5.5] | 3.8 [2.9,4.7] | 3.5 [2.7,4.4] | 3.2 [2.5,4.1] | <0.001 |
| **SOAW,n(%)** |  |  |  |  |  | <0.001 |
| Spring | 11354 (22.0) | 2668 (20.5) | 2796 (21.7) | 2876 (22.3) | 3014 (23.3) |  |
| Summer | 13470 (26.0) | 3276 (25.2) | 3307 (25.6) | 3322 (25.7) | 3565 (27.6) |  |
| Autumn | 15784 (30.5) | 3981 (30.0) | 3867 (30.0) | 4025 (31.2) | 3911 (30.3) |  |
| Winter | 11109 (21.5) | 3065 (23.6) | 2923 (22.7) | 2684 (20.8) | 2437 (18.9) |  |
| **Hypertension,n(%)** |  |  |  |  |  | <0.001 |
| No | 27140 (52.5) | 5904 (45.5) | 6568 (50.9) | 7112 (55.1) | 7556 (58.5) |  |
| Yes | 24577 (47.5) | 7086 (54.5) | 6325 (49.1) | 5795 (44.9) | 5371 (41.5) |  |
| **Platelet** | 250 [210,280] | 250 [210,290] | 250 [210,280] | 250 [210,280] | 240 [210,280] | <0.001 |
| **ALT** | 20 [15,27] | 21 [16,28] | 20 [15,27] | 19 [15,26] | 19 [15,25] | <0.001 |
| **AST** | 24 [21,29] | 24 [21,29] | 24 [21,28] | 24 [21,28] | 25 [21,29] | <0.001 |
| **cholesterol** | 5.7 [5.0,6.4] | 5.6 [4.9,6.4] | 5.7 [5.0,6.5] | 5.7 [5.0,6.4] | 5.6 [5.0,6.4] | <0.001 |
| **Triglycerides** | 1.4 [1.0,2.0] | 1.6 [1.1,2.3] | 1.5 [1.0,2.1] | 1.4 [0.98,2.0] | 1.3 [0.90,1.8] | <0.001 |
| **Wear duration overall** | 6.9 [6.7,7.0] | 6.9 [6.6,7.0] | 6.9 [6.7,7.0] | 6.9 [6.7,7.0] | 6.9 [6.7,7.0] | <0.001 |
| **n-3 PUFA (%TFA)** | 4.3 [3.4,5.3] | 4.3 [3.5,5.3] | 4.3 [3.5,5.3] | 4.3 [3.5,5.3] | 4.2 [3.4,5.2] | <0.001 |
| **n-6 PUFA (%TFA)** | 39 [36,41] | 38 [35,40] | 39 [36,41] | 39 [37,41] | 40 [37,41] | <0.001 |
| **MUFA (%TFA)** | 23 [22,25] | 24 [22,26] | 23 [22,25] | 23 [21,25] | 23 [21,24] | <0.001 |
| **SFA (%TFA)** | 34 [33,35] | 34 [33,35] | 34 [33,35] | 34 [33,35] | 34 [33,35] | <0.001 |

Values are the median (IQR) or percentage (%) and were examined by Kruskal-Wallis or chi-square test;TDI,Townsend deprivation index;SAOW,Season of accelerometer wear.

### 3.2 MVPA and fatty acids’ independent associations with the risk of MASLD

Fig. 2 (Supplementary Tables 1–2) presents the independent associations of MVPA and fatty acids with MASLD risk. After full adjustment, compared to the lowest quartile (Q1), the highest quartile (Q4) of MVPA was associated with a 59% reduced risk (HR = 0.41, 95% CI: 0.30–0.56, *P* < 0.001), and n-6 PUFA Q4 with a 58% reduced risk (HR = 0.42, 95% CI: 0.29–0.62, *P* < 0.001). No significant association was observed for n-3 PUFA. In contrast, MUFA Q4 was associated with a 59% increased risk (HR = 1.59, 95% CI: 1.10–2.30, *P* = 0.014), and SFA Q4 with an 89% increased risk (HR = 1.89, 95% CI: 1.41–2.54, *P* < 0.001). Kaplan-Meier curves (Fig. 3) showed that the highest crude incidence rates were observed in the lowest MVPA and n-6 PUFA levels, and in the highest MUFA and SFA levels, with all differences statistically significant.

**Fig. 2.**
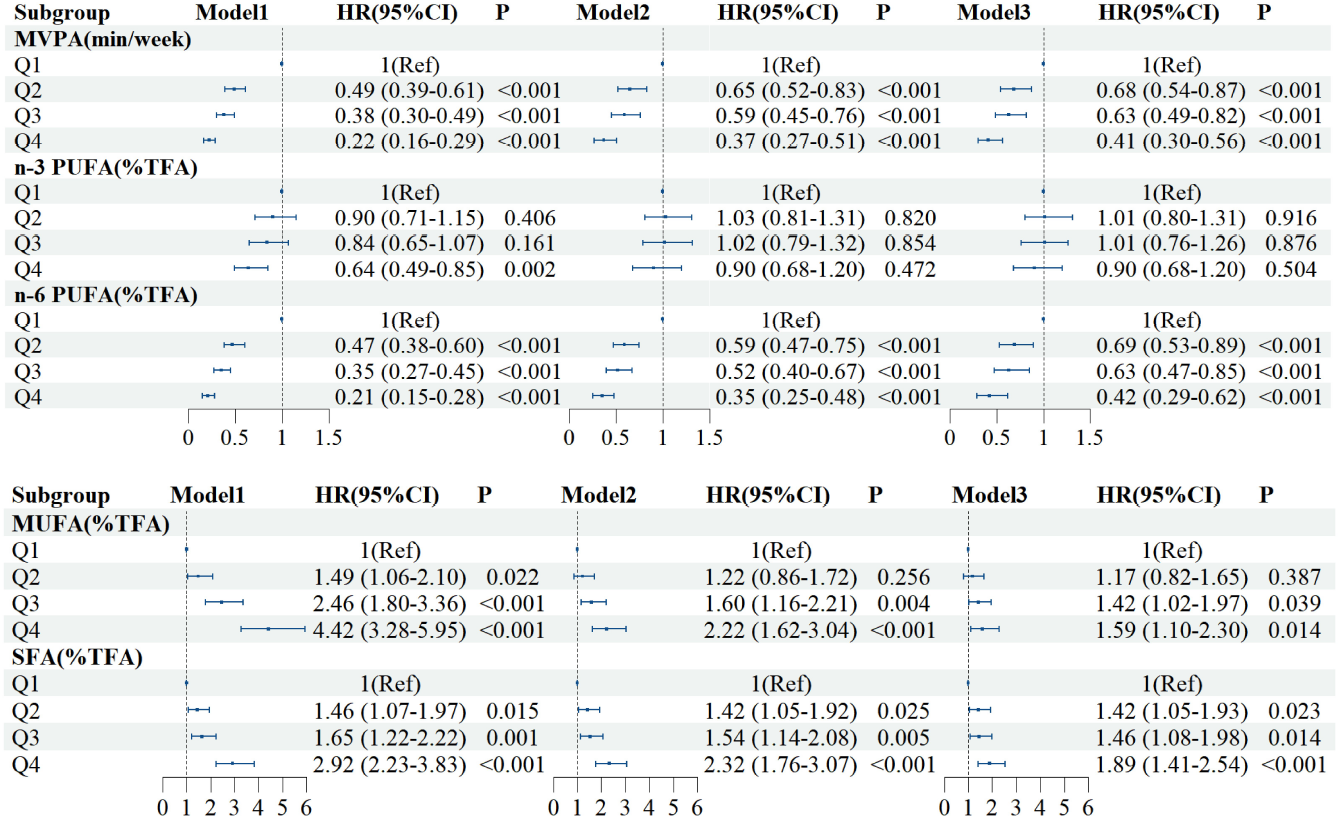
Associations between MVPA,plasma n-3 PUFA, n-6 PUFA, MUFA, SFA levels and incident MASLD risk (n = 51,717 participants). TMultivariable Cox proportional hazard models were used: Model 1 was adjusted for age, sex, and ethnicity. Model 2 was adjusted for Model 1 plus Townsend deprivation index, assessment centre, education, sleep duration, smoking status, alcohol consumption, BMI, and season of accelerometer wearing.Model 3 was adjusted for Model 2 plus hypertension, platelet count, ALT, AST, cholesterol, and triglycerides.

**Fig. 3.**
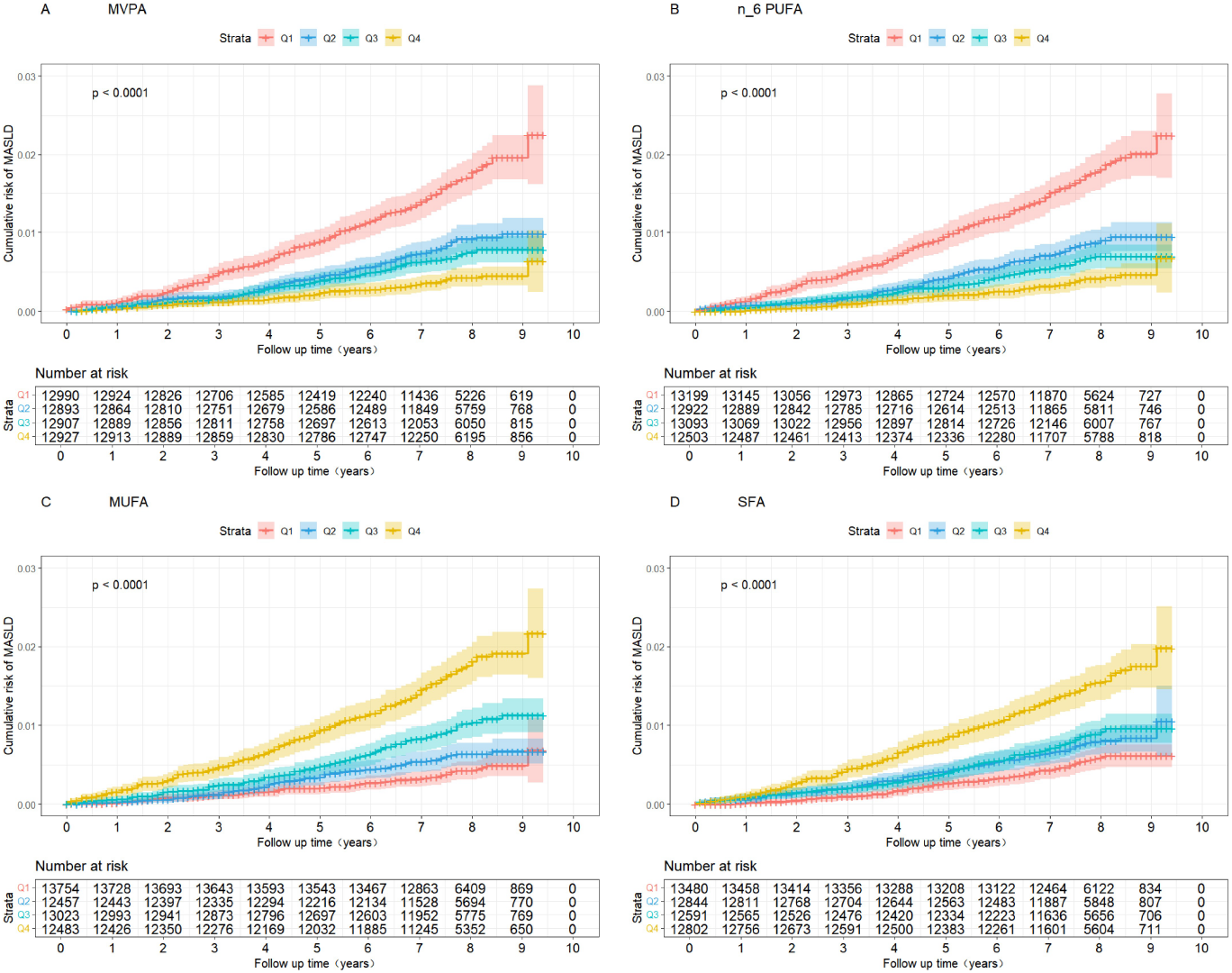
(A) Kaplan-Meier survival estimates according to MVPA level for the probability of incident MASLD.(B) Kaplan-Meier survival estimates according to n-6 PUFA level for the probability of incident MASLD.(C) Kaplan-Meier survival estimates according to MUFA level for the probability of incident MASLD. (D) Kaplan-Meier survival estimates according to SFA level for the probability of incident MASLD.

### 3.3 The dose-response associations of MVPA and fatty acids with the risk of MASLD

To assess linear or nonlinear associations, multivariable-adjusted restricted cubic spline (RCS) analysis was performed (Fig. 4). MUFA and SFA showed linear positive associations with MASLD risk (nonlinear *P* > 0.05), while n-6 PUFA showed a linear negative association (nonlinear *P* > 0.05). MVPA exhibited a nonlinear negative relationship (nonlinear *P* < 0.05). Threshold effect analysis using a piecewise Cox model identified an inflection point at 189 (likelihood ratio test *P* < 0.05; Supplementary Table 3). On both sides of this threshold, higher MVPA was significantly associated with reduced MASLD risk.

**Fig. 4.**
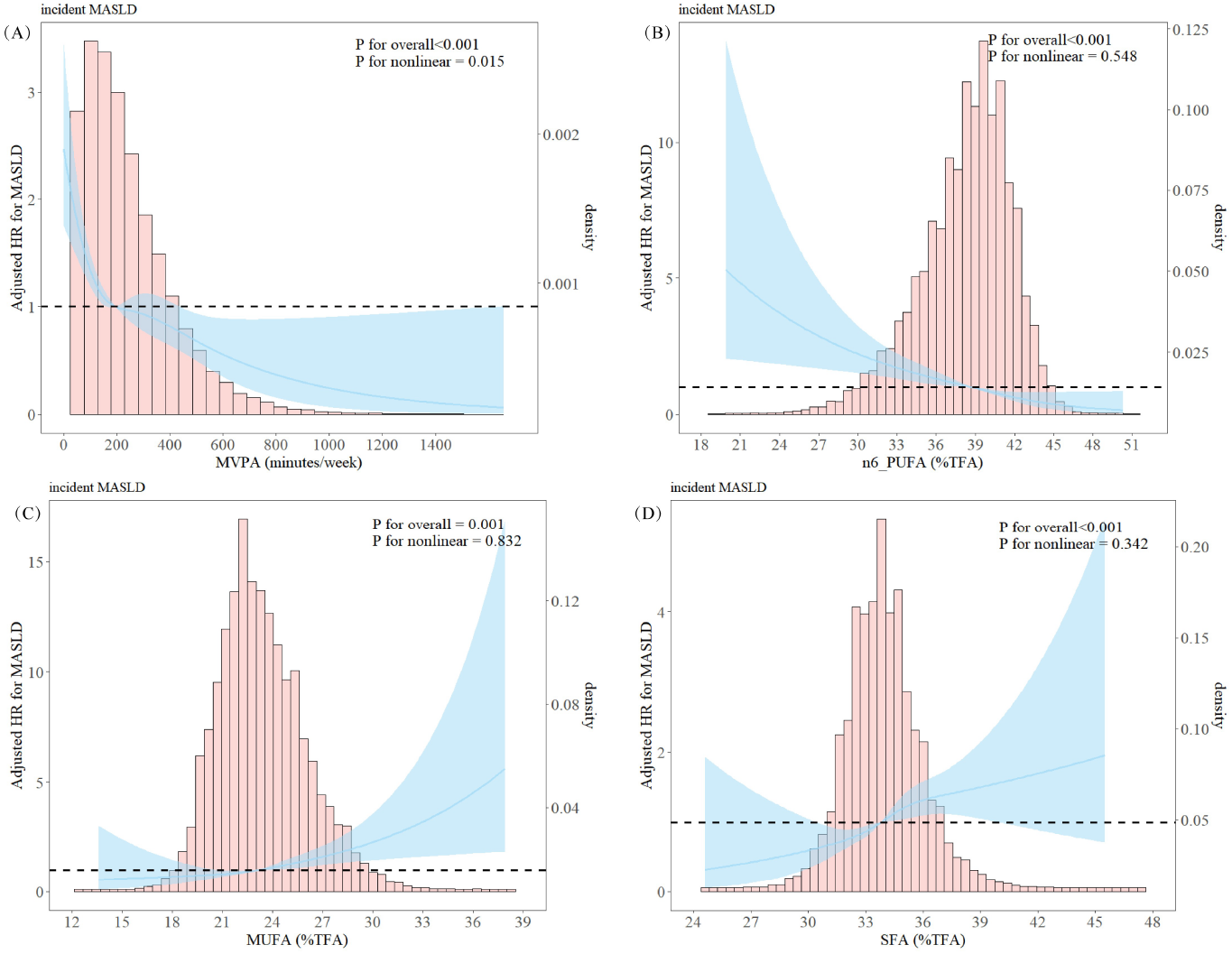
The dose-response relationship between MVPA, different fatty acids, and the risk of MASLD. The histogram shows the distribution of different exposure levels in the overall population. The results have been adjusted for age, sex, ethnicity, Townsend deprivation index, assessment centre, education, sleep duration, smoking status, alcohol consumption, BMI, season of accelerometer wearing, hypertension, platelet count, ALT, AST, cholesterol, and triglycerides.

### 3.4 The joint association of MVPA and fatty acids with the risk of MASLD

Supplementary figure 5 presents the joint analysis of MVPA and fatty acids with MASLD risk. Compared to *Not Recommended* MVPA combined with n-6 PUFA Q1, the combination of *Recommended* MVPA and n-6 PUFA Q4 showed the lowest risk (HR = 0.29, 95% CI: 0.18–0.45). Compared to *Not Recommended* MVPA with MUFA Q4, *Recommended* MVPA with MUFA Q1 had the lowest risk (HR = 0.39, 95% CI: 0.25–0.62). Compared to *Not Recommended* MVPA with SFA Q4, *Recommended* MVPA with SFA Q1 had the lowest risk (HR = 0.37, 95% CI: 0.25–0.55). Within each MVPA category, the lowest risk was observed in n-6 PUFA Q4 and SFA Q1, except for the *Not Recommended* MVPA with MUFA combination, which was not statistically significant.

### 3.5 Subgroup and sensitivity analyses

Supplementary tables 4-7 present the results of subgroup analyses stratified by sex, age, ethnicity, recruitment centre, education, smoking status, alcohol consumption, BMI, season of accelerometer wearing, and hypertension. Notably, in the subgroup analysis of the association between MVPA and MASLD, ethnicity interacted with the risk of MASLD (interaction P < 0.05). In the subgroup analysis of the association between SFA and MASLD, smoking status interacted with the risk of MASLD (interaction P < 0.05), but both subgroups were closely associated with MASLD. However, the interaction between sex, age, recruitment centre, education, alcohol consumption, BMI, season of accelerometer wearing, and hypertension with the risk of MASLD was not significant. This suggests that the effects of MVPA and fatty acids on the risk of MASLD are consistent across different demographic and clinical subgroups. To validate the robustness of the primary findings, several sensitivity analyses were conducted. First, a sensitivity analysis that excluded participants with follow-up within 2 years demonstrated results consistent with the primary analysis, thereby minimising potential reverse causation. Second, the complete-case dataset excluding participants with missing covariates showed concordant results with the primary analysis. The third step excluded participants with hypertension and diabetes at baseline, but it did not significantly alter our research results(Supplementary Tables 8-10).

## 4 Discussion

This prospective cohort study examined the individual and joint effects of accelerometer-measured MVPA and plasma fatty acids on MASLD risk. MVPA and n-6 PUFA were associated with reduced risk, with a nonlinear dose-response relationship for MVPA, while n-3 PUFA showed no significant association. MUFA and SFA were associated with increased risk. In joint analyses, the lowest risk was observed for Recommended MVPA combined with n-6 PUFA Q4 (HR = 0.29, 95% CI: 0.18–0.45), MUFA Q1 (HR = 0.39, 95% CI: 0.25–0.62), or SFA Q1 (HR = 0.37, 95% CI: 0.25–0.55). These findings remained robust in sensitivity and subgroup analyses. Few studies have explored the joint effects of MVPA and fatty acids on MASLD, and this study adds prospective evidence to the literature.

To date, only two studies have reported a negative association between accelerometer-measured physical activity (PA) and MASLD risk (Schneider et al. 2021; Liu et al. 2024). Our findings align with these, providing further evidence using short-duration MVPA captured by a wrist-worn accelerometer (Li et al. 2025).Regarding fatty acids, previous studies have shown that saturated fatty acids (SFA) are positively associated with liver disease risk, while n-3 and n-6 polyunsaturated fatty acids (PUFA) are protective (Liu et al. 2024; Kaikkonen et al. 2017). However, our study found that n-6 PUFA, but not n-3 PUFA, was associated with reduced MASLD risk. This finding is consistent with a recent Serbian study (Frankovic et al. 2024), suggesting that n-6 PUFA may play a particularly important role in MASLD development.

Few studies have examined the joint effects of MVPA and fatty acids on MASLD risk. A US prospective study found that MVPA was positively correlated with individual MUFAs during late pregnancy, suggesting MVPA may influence plasma MUFA levels (Xia et al. 2022). Exercise is known to improve MASLD by enhancing fatty acid *β*-oxidation, inducing liver-protective autophagy, and increasing insulin sensitivity (Farzanegi et al. 2019). While n-3 PUFA is thought to protect against liver disease via anti-inflammatory effects (de Castro and Calder 2018), the role of n-6 PUFA in inflammation remains debated (Tortosa-Caparrós et al. 2017). Notably, our study found n-6 PUFA, but not n-3 PUFA, was associated with reduced MASLD risk.

This study is the first to explore the joint effects of MVPA and fatty acids on MASLD risk. Despite its large sample size, long follow-up, and objective measurements, several limitations remain. First, accelerometer wear began years after baseline, introducing a potential time gap, though previous studies indicate moderate-to-high reproducibility of activity measurements over time (Pernar et al. 2022; Al-Shaar et al. 2022; Keadle et al. 2017). Second, plasma fatty acids were measured only once at baseline, which may not reflect long-term exposure. Third, residual confounding cannot be ruled out due to the observational design. Finally, UKB participants are not fully representative of the general UK population, with a low response rate and overrepresentation of healthier, white European individuals (Fry et al. 2017), which may limit generalisability. Nonetheless, the large-scale exposure data provide valuable scientific insights (Batty et al. 2020).

## 5 Conclusion

The findings from the UKB study indicate a significant association between MVPA and specific fatty acid levels and the risk of MASLD. Specifically, higher levels of MVPA and plasma n-6 PUFA are associated with a reduced risk of MASLD. In comparison, higher levels of MUFA and SFA are associated with an increased risk of MASLD. Notably, an increase in plasma n-3 PUFA was not associated with a reduced risk of MASLD. The joint analysis of MVPA and fatty acids reveals that combining recommended MVPA with specific levels of n-6 PUFA, MUFA, and SFA significantly reduces the risk of MASLD. Our study contributes prospective evidence on the effects of MVPA and fatty acids on MASLD risk, providing theoretical support for developing personalised health interventions. This suggests that, in preventing MASLD, maintaining appropriate levels of physical activity and adjusting fatty acid levels may be important.

## Data Availability

The datasets analysed during the current study are available from the UK Biobank through an application process (https://www.ukbiobank.ac.uk/). This study was conducted using the UK Biobank Resource under Application Number 160957.

## Abbreviations

ALT: Alanine aminotransferase
AST: Aspartate aminotransferase
BMI: Body mass index
CI: Confidence interval
HR: Hazard ratio
ICD-10: International Classification of Diseases
MASLD: Metabolic dysfunction-associated steatotic liver disease
MVPA: Moderate-to-vigorous physical activity
MUFA: Monounsaturated fatty acids
PA: Physical activity
PUFA: Polyunsaturated fatty acids
SFA: Saturated fatty acids
TDI: Townsend deprivation index
WHO: World health organization

## Supplementary information

Table S1. Associations between MVPA, plasma n-3 PUFA, n-6 PUFA levels and incident MASLD risk. Table S2. Associations between plasma MUFA, SFA levels and incident MASLD risk. Table S3. Threshold effect analysis of MVPA on the risk of MASLD incidence using a piecewise regression model. Table S4. HR (95%CI) for incident MASLD associated with MVPA in various subgroups. Table S5. HR (95%CI) for incident MASLD associated with plasma n-6 PUFA in various subgroups. Table S6. HR (95%CI) for incident MASLD associated with plasma MUFA in various subgroups. Table S7. HR (95%CI) for incident MASLD associated with plasma SFA in various subgroups. Table S8. Sensitivity analysis of the associations between MVPA,plasma n-6 PUFA,MUFA,SFA levels and incident MASLD risk by excluding participants within 2 years of follow-up. Table S9. Sensitivity analysis of the associations between MVPA,plasma n-6 PUFA,MUFA,SFA levels and incident MASLD risk using a complete-case dataset without missing values. Table S10. Sensitivity analysis of the associations between MVPA,plasma n-6 PUFA,MUFA,SFA levels and incident MASLD risk by excluding participants with hypertension or diabetes at baseline. Fig S1. The joint association of MVPA and fatty acids with the risk of MASLD.

## Declarations

### Ethics approval and consent to participate

The North West Research Ethics Committee approved the UKB (21/NW/0157) and all participants signed an informed consent .

### Consent for publication

Not applicable.

### Availability of data and Materials

The datasets analysed during the current study are available from the UKB through an application process (http://www.ukbiobank.ac.uk/).

## References

Arnett, D.K., Blumenthal, R.S., Albert, M.A., Buroker, A.B., Goldberger, Z.D., Hahn, E.J., Himmelfarb, C.D., Khera, A., Lloyd-Jones, D., McEvoy, J.W., et al.: Correction to: 2019 acc/aha guideline on the primary prevention of cardiovascular disease: a report of the american college of cardiology/american heart association task force on clinical practice guidelines. Circulation 140(11), 649–650 (2019)

Al-Shaar, L., Pernar, C.H., Chomistek, A.K., Rimm, E.B., Rood, J., Stampfer, M.J., Eliassen, A.H., Barnett, J.B., Willett, W.C.: Reproducibility, validity, and relative validity of self-report methods for assessing physical activity in epidemiologic studies: findings from the women’s lifestyle validation study. American journal of epidemiology 191(4), 696–710 (2022)

Bull, F.C., Al-Ansari, S.S., Biddle, S., Borodulin, K., Buman, M.P., Cardon, G., Carty, C., Chaput, J.-P., Chastin, S., Chou, R., et al.: World health organization 2020 guidelines on physical activity and sedentary behaviour. British journal of sports medicine 54(24), 1451–1462 (2020)

Bansal, S.K., Bansal, M.B.: Pathogenesis of masld and mash–role of insulin resistance and lipotoxicity. Alimentary Pharmacology & Therapeutics 59, 10–22 (2024)

Batty, G.D., Gale, C.R., Kivimäki, M., Deary, I.J., Bell, S.: Comparison of risk factor associations in uk biobank against representative, general population based studies with conventional response rates: prospective cohort study and individual participant meta-analysis. bmj 368 (2020)

Chan, W.-K., Chuah, K.-H., Rajaram, R.B., Lim, L.-L., Ratnasingam, J., Vethakkan, S.R.: Metabolic dysfunction-associated steatotic liver disease (masld): a state-of-the-art review. Journal of obesity & metabolic syndrome 32(3), 197 (2023)

Cui, J., Li, L., Ren, L., Sun, J., Zhao, H., Sun, Y., Cui, J., Li, L., Ren, L., Sun, J., et al.: Dietary n-3 and n-6 fatty acid intakes and nafld: A cross-sectional study in the united states. Asia Pacific journal of clinical nutrition 30(1) (2021)

DiPietro, L., Al-Ansari, S.S., Biddle, S.J., Borodulin, K., Bull, F.C., Buman, M.P., Cardon, G., Carty, C., Chaput, J.-P., Chastin, S., et al.: Advancing the global physical activity agenda: recommendations for future research by the 2020 who physical activity and sedentary behavior guidelines development group. International Journal of Behavioral Nutrition and Physical Activity 17, 1–11 (2020)

Castro, G.S., Calder, P.C.: Non-alcoholic fatty liver disease and its treatment with n-3 polyunsaturated fatty acids. Clinical nutrition 37(1), 37–55 (2018)

Doherty, A., Jackson, D., Hammerla, N., Plötz, T., Olivier, P., Granat, M.H., White, T., Van Hees, V.T., Trenell, M.I., Owen, C.G., et al.: Large scale population assessment of physical activity using wrist worn accelerometers: the uk biobank study. PloS one 12(2), 0169649 (2017)

Farzanegi, P., Dana, A., Ebrahimpoor, Z., Asadi, M., Azarbayjani, M.A.: Mechanisms of beneficial effects of exercise training on non-alcoholic fatty liver disease (nafld): Roles of oxidative stress and inflammation. European journal of sport science 19(7), 994–1003 (2019)

Frankovic, I., Djuricic, I., Ninic, A., Vekic, J., Vorkapic, T., Erceg, S., Gojkovic, T., Tomasevic, R., Mamic, M., Mitrovic, M., et al.: Increased odds of metabolic dysfunction-associated steatotic liver disease are linked to reduced n-6, but not n-3 polyunsaturated fatty acids in plasma. Biomolecules 14(8), 902 (2024)

Füzéki, E., Engeroff, T., Banzer, W.: Health benefits of light-intensity physical activity: a systematic review of accelerometer data of the national health and nutrition examination survey (nhanes). Sports medicine 47, 1769–1793 (2017)

Fry, A., Littlejohns, T.J., Sudlow, C., Doherty, N., Adamska, L., Sprosen, T., Collins, R., Allen, N.E.: Comparison of sociodemographic and health-related characteristics of uk biobank participants with those of the general population. American journal of epidemiology 186(9), 1026–1034 (2017)

Feng, H., Yang, L., Liang, Y.Y., Ai, S., Liu, Y., Liu, Y., Jin, X., Lei, B., Wang, J., Zheng, N., et al.: Associations of timing of physical activity with all-cause and cause-specific mortality in a prospective cohort study. Nature communications 14(1), 930 (2023)

Julkunen, H., Cichońska, A., Tiainen, M., Koskela, H., Nybo, K., Mäkelä, V., Nokso-Koivisto, J., Kristiansson, K., Perola, M., Salomaa, V., et al.: Atlas of plasma nmr biomarkers for health and disease in 118,461 individuals from the uk biobank. Nature communications 14(1), 604 (2023)

Jones, S.M., Porroche-Escudero, A., Shearn, K., Hunter, R.F., Garcia, L.: Thinking about inequalities in physical activity as an emergent feature of complex systems. International Journal of Behavioral Nutrition and Physical Activity 21(1), 125 (2024)

Keadle, S.K., Shiroma, E.J., Kamada, M., Matthews, C.E., Harris, T.B., Lee, I.-M.: Reproducibility of accelerometer-assessed physical activity and sedentary time. American journal of preventive medicine 52(4), 541–548 (2017)

Khurshid, S., Weng, L.-C., Al-Alusi, M.A., Halford, J.L., Haimovich, J.S., Benjamin, E.J., Trinquart, L., Ellinor, P.T., McManus, D.D., Lubitz, S.A.: Accelerometer-derived physical activity and risk of atrial fibrillation. European heart journal 42(25), 2472–2483 (2021)

Kaikkonen, J.E., Würtz, P., Suomela, E., Lehtovirta, M., Kangas, A.J., Jula, A., Mikkilä, V., Viikari, J.S., Juonala, M., Rönnemaa, T., et al.: Metabolic profiling of fatty liver in young and middle-aged adults: Cross-sectional and prospective analyses of the young finns study. Hepatology 65(2), 491–500 (2017)

Li, Z., Cheng, S., Guo, B., Ding, L., Liang, Y., Shen, Y., Li, J., Hu, Y., Long, T., Guo, X., et al.: Wearable device-measured moderate to vigorous physical activity and risk of degenerative aortic valve stenosis. European Heart Journal 46(7), 649–664 (2025)

Liu, Z., Huang, H., Xie, J., Xu, Y., Xu, C.: Circulating fatty acids and risk of hepatocellular carcinoma and chronic liver disease mortality in the uk biobank. Nature Communications 15(1), 3707 (2024)

Lee, W.-H., Kipp, Z.A., Bates, E.A., Pauss, S.N., Martinez, G.J., Hinds Jr, T.D.: The physiology of masld: molecular pathways between liver and adipose tissues. Clinical Science 139(18), 1015–1046 (2025)

Li, P., Yang, Q., Wang, X., Sun, S., Cao, W., Yu, S., Zhan, S., Sun, F.: Dose-response relationship between physical activity and nonalcoholic fatty liver disease: A prospective cohort study. Chinese Medical Journal 136(12), 1494–1496 (2023)

Liu, M., Ye, Z., Zhang, Y., He, P., Zhou, C., Yang, S., Zhang, Y., Gan, X., Qin, X.: Accelerometer-derived moderate-to-vigorous physical activity and incident nonalcoholic fatty liver disease. BMC medicine 22(1), 398 (2024)

Machado, M.V.: Masld treatment—a shift in the paradigm is imminent. Frontiers in medicine 10, 1316284 (2023)

Mudd, A.L., Bal, M., Verra, S.E., Poelman, M.P., Wit, J., Kamphuis, C.B.: The current state of complex systems research on socioeconomic inequalities in health and health behavior—a systematic scoping review. International Journal of Behavioral Nutrition and Physical Activity 21(1), 13 (2024)

Masoodi, M., Gastaldelli, A., Hyötyläinen, T., Arretxe, E., Alonso, C., Gaggini, M., Brosnan, J., Anstee, Q.M., Millet, O., Ortiz, P., et al.: Metabolomics and lipidomics in nafld: biomarkers and non-invasive diagnostic tests. Nature reviews Gastroenterology & hepatology 18(12), 835–856 (2021)

Miao, L., Targher, G., Byrne, C.D., Cao, Y.-Y., Zheng, M.-H.: Current status and future trends of the global burden of masld. Trends in Endocrinology & Metabolism (2024)

Pernar, C.H., Chomistek, A.K., Barnett, J.B., Ivey, K., Al-Shaar, L., Roberts, S.B., Rood, J., Fielding, R.A., Block, J., Li, R., et al.: Validity and relative validity of alternative methods of assessing physical activity in epidemiologic studies: Findings from the men’s lifestyle validation study. American journal of epidemiology 191(7), 1307–1322 (2022)

Qiu, S., Cai, X., Sun, Z., Li, L., Zügel, M., Steinacker, J.M., Schumann, U.: Association between physical activity and risk of nonalcoholic fatty liver disease: a meta-analysis. Therapeutic advances in gastroenterology 10(9), 701–713 (2017)

Sheka, A.C., Adeyi, O., Thompson, J., Hameed, B., Crawford, P.A., Ikramuddin, S.: Nonalcoholic steatohepatitis: a review. Jama 323(12), 1175–1183 (2020)

Summerhayes, L., Baker, D., Vella, K.: Food diversity and accessibility enabled urban environments for sustainable food consumption: a case study of brisbane, australia. Humanities and Social Sciences Communications 11(1), 1–14 (2024)

Schneider, C.V., Zandvakili, I., Thaiss, C.A., Schneider, K.M.: Physical activity is associated with reduced risk of liver disease in the prospective uk biobank cohort. JHEP Reports 3(3), 100263 (2021)

Tortosa-Caparrós, E., Navas-Carrillo, D., Marín, F., Orenes-Piñero, E.: Anti-inflammatory effects of omega 3 and omega 6 polyunsaturated fatty acids in cardiovascular disease and metabolic syndrome. Critical reviews in food science and nutrition 57(16), 3421–3429 (2017)

Truong, X.T., Lee, D.H.: Hepatic insulin resistance and steatosis in metabolic dysfunction-associated steatotic liver disease: new insights into mechanisms and clinical implications. Diabetes & Metabolism Journal 49(5), 964–986 (2025)

Van Der Windt, D.J., Sud, V., Zhang, H., Tsung, A., Huang, H.: The effects of physical exercise on fatty liver disease. Gene expression 18(2), 89 (2018)

Wang, D., Dai, X., Mishra, S.R., Lim, C.C., Carrillo-Larco, R.M., Gakidou, E., Xu, X.: Association between socioeconomic status and health behaviour change before and after non-communicable disease diagnoses: a multicohort study. The Lancet Public Health 7(8), 670–682 (2022)

White, T., Westgate, K., Hollidge, S., Venables, M., Olivier, P., Wareham, N., Brage, S.: Estimating energy expenditure from wrist and thigh accelerometry in free-living adults: a doubly labelled water study. International journal of obesity 43(11), 2333–2342 (2019)

Xia, T., Chen, L., Fei, Z., Liu, X., Dai, J., Hinkle, S.N., Zhu, Y., Wu, J., Weir, N.L., Tsai, M.Y., et al.: A longitudinal study on associations of moderate-to-vigorous physical activity with plasma monounsaturated fatty acids in pregnancy. Frontiers in Nutrition 9, 983418 (2022)

