## Supplementary information for "Physical activity, fatty acids, and MASLD risk: Behavioural and metabolic factors jointly shaping liver health in populations"

**Supplemental Materials**

**Table S1**. Associations between MVPA, plasma n-3 PUFA, n-6 PUFA levels and incident MASLD risk.

**Table S2**. Associations between plasma MUFA, SFA levels and incident MASLD risk.

**Table S3**. Threshold effect analysis of MVPA on the risk of MASLD incidence using a piecewise regression model.

**Table S4**. HR (95%CI) for incident MASLD associated with MVPA in various subgroups.

**Table S5**. HR (95%CI) for incident MASLD associated with plasma n-6 PUFA in various subgroups.

**Table S6**. HR (95%CI) for incident MASLD associated with plasma MUFA in various subgroups.

**Table S7**. HR (95%CI) for incident MASLD associated with plasma SFA in various subgroups.

**Table S8**. Sensitivity analysis of the associations between MVPA，plasma n-6 PUFA，MUFA，SFA levels and incident MASLD risk by excluding participants within 2 years of follow-up.

**Table S9**. Sensitivity analysis of the associations between MVPA，plasma n-6 PUFA，MUFA，SFA levels and incident MASLD risk using a complete-case dataset without missing values.

**Table S10**. Sensitivity analysis of the associations between MVPA，plasma n-6 PUFA，MUFA，SFA levels and incident MASLD risk by excluding participants with hypertension or diabetes at baseline.

**Fig S1.** The joint association of MVPA and fatty acids with the risk of MASLD.

**Table S1. Associations between MVPA, plasma n-3 PUFA, n-6 PUFA levels and incident MASLD risk.**

| **Exposure Group** | **Events/n(rate^a^)** | **Model 1** |  | **Model 2** |  | **Model 3** |  |
| --- | --- | --- | --- | --- | --- | --- | --- |
|  |  | **HR (95% CI)** | ***P* value** | **HR (95% CI)** | ***P* value** | **HR (95% CI)** | ***P* value** |
| **MVPA(min/week)** |  |  |  |  |  |  |  |
| **Q1** | 214/12990(2.1) | 1(Ref) |  | 1(Ref) |  | 1(Ref) |  |
| **Q2** | 112/12893(1.1) | 0.49 (0.39-0.61) | <0.001 | 0.65 (0.52-0.83) | <0.001 | 0.68 (0.54-0.87) | <0.001 |
| **Q3** | 92/12907(0.9) | 0.38 (0.30-0.49) | <0.001 | 0.59 (0.45-0.76) | <0.001 | 0.63 (0.49-0.82) | <0.001 |
| **Q4** | 54/12927(0.5) | 0.22 (0.16-0.29) | <0.001 | 0.37 (0.27-0.51) | <0.001 | 0.41 (0.30-0.56) | <0.001 |
| **n-3 PUFA(%TFA)** |  |  |  |  |  |  |  |
| **Q1** | 142/13183(1.4) | 1(Ref) |  | 1(Ref) |  | 1(Ref) |  |
| **Q2** | 133/13691(1.3) | 0.90 (0.71-1.15) | 0.406 | 1.03 (0.81-1.31) | 0.82 | 1.01 (0.80-1.31) | 0.916 |
| **Q3** | 112/12484(1.2) | 0.84 (0.65-1.07) | 0.161 | 1.02 (0.79-1.32) | 0.854 | 1.01 (0.76-1.26) | 0.876 |
| **Q4** | 85/12359(0.9) | 0.64 (0.49-0.85) | 0.002 | 0.90 (0.68-1.20) | 0.472 | 0.90 (0.68-1.20) | 0.504 |
| **n-6 PUFA(%TFA)** |  |  |  |  |  |  |  |
| **Q1** | 228/13199(2.3) | 1(Ref) |  | 1(Ref) |  | 1(Ref) |  |
| **Q2** | 109/12922(1.1) | 0.47 (0.38-0.60) | <0.001 | 0.59 (0.47-0.75) | <0.001 | 0.69 (0.53-0.89) | <0.001 |
| **Q3** | 84/13093(0.8) | 0.35 (0.27-0.45) | <0.001 | 0.52 (0.40-0.67) | <0.001 | 0.63 (0.47-0.85) | <0.001 |
| **Q4** | 51/12503(0.5) | 0.21 (0.15-0.28) | <0.001 | 0.35 (0.25-0.48) | <0.001 | 0.42 (0.29-0.62) | <0.001 |

^a^Incidence rates were expressed as per 1000 person-years

Multivariable Cox proportional hazard models were used:Model 1 was adjusted for age, sex, and ethnicity.Model 2 was adjusted for Model 1 plus Townsend deprivation index, assessment centre, education, sleep duration, smoking status,alcohol consumption, BMI, and season of accelerometer wearing.Model 3 was adjusted for Model 2 plus hypertension, platelet count, ALT, AST, cholesterol, and triglycerides.

**Table S2. Associations between plasma MUFA, SFA levels and incident MASLD risk.**

| **Exposure Group** | **Events/n(rate^a^)** | **Model 1** |  | **Model 2** |  | **Model 3** |  |
| --- | --- | --- | --- | --- | --- | --- | --- |
|  |  | **HR (95% CI)** | ***P* value** | **HR (95% CI)** | ***P* value** | **HR (95% CI)** | ***P* value** |
| **MUFA(%TFA)** |  |  |  |  |  |  |  |
| **Q1** | 58/13754(0.5) | 1(Ref) |  | 1(Ref) |  | 1(Ref) |  |
| **Q2** | 76/12457(0.8) | 1.49 (1.06-2.10) | 0.022 | 1.22 (0.86-1.72) | 0.256 | 1.17 (0.82-1.65) | 0.387 |
| **Q3** | 127/13023(1.3) | 2.46 (1.80-3.36) | <0.001 | 1.60 (1.16-2.21) | 0.004 | 1.42 (1.02-1.97) | 0.039 |
| **Q4** | 211/12483(2.2) | 4.42 (3.28-5.95) | <0.001 | 2.22 (1.62-3.04) | <0.001 | 1.59 (1.10-2.30) | 0.014 |
| **SFA(%TFA)** |  |  |  |  |  |  |  |
| **Q1** | 73/13480(0.7) | 1(Ref) |  | 1(Ref) |  | 1(Ref) |  |
| **Q2** | 98/12844(1.0) | 1.46 (1.07-1.97) | 0.015 | 1.42 (1.05-1.92) | 0.025 | 1.42 (1.05-1.93) | 0.023 |
| **Q3** | 108/12591(1.1) | 1.65 (1.22-2.22) | 0.001 | 1.54 (1.14-2.08) | 0.005 | 1.46 (1.08-1.98) | 0.014 |
| **Q4** | 193/12802(2.0) | 2.92 (2.23-3.83) | <0.001 | 2.32 (1.76-3.07) | <0.001 | 1.89 (1.41-2.54) | <0.001 |

^a^Incidence rates were expressed as per 1000 person-years

Multivariable Cox proportional hazard models were used:Model 1 was adjusted for age, sex, and ethnicity.Model 2 was adjusted for Model 1 plus Townsend deprivation index, assessment centre, education, sleep duration, smoking status,alcohol consumption, BMI, and season of accelerometer wearing.Model 3 was adjusted for Model 2 plus hypertension, platelet count, ALT, AST, cholesterol, and triglycerides.

**Table S3.Threshold effect analysis of MVPA on the risk of MASLD incidence using a piecewise regression model.**

| **The effect siz** | **Adjusted HR (95%CI)** | **P value** |
| --- | --- | --- |
| **MVPA** |  |  |
| **Total** | 0.996(0.995-0.997) | <0.001 |
| **Fitting model by two-piecewise**  **Cox proportional risk model** |  |  |
| **Inflection point** | 189 |  |
| **<189** | 0.993(0.991-0.994) | <0.001 |
| **>189** | 0.998(0.997-1.000) | 0.006 |
| **P for log-likelihood ratio** |  | <0.001 |

Multivariable Cox proportional hazard models were used.Model was adjusted for age, sex, ethnicity, townsend deprivation index, assessment centre, education, sleep duration, smoking status,alcohol consumption, BMI, season of accelerometer wearing, hypertension, platelet count, ALT, AST, cholesterol, and triglycerides.

**Table S4. HR (95%CI) for incident MASLD associated with MVPA in various subgroups.**

| **Subgroups** | **Events/n(%)** | | **MVPA Q2 (108-196)** | | **MVPA Q3 (196-320)** | | **MVPA Q4 (320-2454)** | | ***P* for interaction** | |
| --- | --- | --- | --- | --- | --- | --- | --- | --- | --- | --- |
| **Sex** |  | |  | |  | |  | | 0.435 | |
| **Female** | 248/28882 (0.9%) | | 0.72 (0.53-0.99) | | 0.55 (0.38-0.79) | | 0.33 (0.20-0.53) | |  | |
| **Male** | 224/22835 (1%) | | 0.64 (0.44-0.91) | | 0.72 (0.50-1.04) | | 0.50 (0.32-0.77) | |  | |
| **Age,years** |  | |  | |  | |  | | 0.880 | |
| **< 60** | 295/30838 (1%) | | 0.75 (0.55-1.01) | | 0.69 (0.50-0.96) | | 0.49 (0.34-0.71) | |  | |
| **≥ 60** | 177/20879 (0.8%) | | 0.58 (0.40-0.85) | | 0.52 (0.33-0.82) | | 0.28 (0.15-0.54) | |  | |
| **Ethnicity** |  | |  | |  | |  | | 0.045 | |
| **Nonwhite** | 22/1454 (1.5%) | | 1.03 (0.33-3.17) | | 0.16 (0.02-1.41) | | 1.69 (0.48-5.94) | |  | |
| **White** | 450/50263 (0.9%) | | 0.66 (0.52-0.84) | | 0.64 (0.49-0.83) | | 0.37 (0.26-0.51) | |  | |
| **Recruitment center** |  | |  | |  | |  | | 0.605 | |
| **England** | 452/47461 (1%) | | 0.67 (0.53-0.85) | | 0.63 (0.48-0.82) | | 0.38 (0.27-0.53) | |  | |
| **Scotland** | 10/2748 (0.4%) | | 0.56 (0.09-3.49) | | 0.38 (0.03-4.82) | | 1.55 (0.22-11.04) | |  | |
| **Wales** | 10/1508 (0.7%) | | 1.02 (0.20-5.38) | | 0.55 (0.05-5.90) | | 1.63 (0.24-11.02) | |  | |
| **education** |  | |  | |  | |  | | 0.962 | |
| **Any other qualification** | 266/25105 (1.1%) | | 0.70 (0.51-0.95) | | 0.63 (0.45-0.90) | | 0.43 (0.28-0.67) | |  | |
| **Degree or above** | 150/22030 (0.7%) | | 0.73 (0.47-1.12) | | 0.68 (0.43-1.06) | | 0.45 (0.27-0.78) | |  | |
| **No qualification** | 56/4582 (1.2%) | | 0.48 (0.23-0.99) | | 0.44 (0.19-1.02) | | 0.14 (0.04-0.53) | |  | |
| **Smoking status** |  | |  | |  | |  | | 0.164 | |
| **Current** | 55/3538 (1.6%) | | 0.86 (0.43-1.69) | | 0.41 (0.16-1.06) | | 1.01 (0.45-2.26) | |  | |
| **Never** | 217/29528 (0.7%) | | 0.75 (0.53-1.07) | | 0.69 (0.47-1.01) | | 0.50 (0.32-0.80) | |  | |
| **Previous** | 200/18651 (1.1%) | | 0.56 (0.39-0.81) | | 0.58 (0.39-0.85) | | 0.25 (0.14-0.43) | |  | |
| **Alcohol consumption** |  | |  | |  | |  | | 0.314 | |
| **<3 times/week** | 238/23539 (1%) | | 0.90 (0.65-1.23) | | 0.66 (0.45-0.96) | | 0.46 (0.29-0.74) | |  | |
| **3+ times/week** | | 186/25297 (0.7%) | | 0.48 (0.32-0.72) | | 0.55 (0.37-0.82) | | 0.39 (0.24-0.62) | |  |
| **Never** | | 48/2881 (1.7%) | | 0.52 (0.23-1.18) | | 0.83 (0.38-1.80) | | 0.29 (0.08-1.03) | |  |
| **BMI,kg/m^2^** | |  | |  | |  | |  | | 0.323 |
| **< 25** | | 61/19957 (0.3%) | | 0.36 (0.17-0.78) | | 0.53 (0.27-1.02) | | 0.36 (0.17-0.74) | |  |
| **≥ 25** | | 411/31760 (1.3%) | | 0.73 (0.57-0.94) | | 0.64 (0.48-0.85) | | 0.43 (0.30-0.62) | |  |
| **Season of accelerometer wearing** | |  | |  | |  | |  | | 0.950 |
| **Autumn** | | 153/15784 (1%) | | 0.64 (0.42-0.97) | | 0.54 (0.34-0.85) | | 0.37 (0.21-0.65) | |  |
| **Spring** | | 83/11354 (0.7%) | | 0.62 (0.34-1.11) | | 0.70 (0.38-1.27) | | 0.39 (0.18-0.85) | |  |
| **Summer** | | 131/13470 (1%) | | 0.59 (0.37-0.93) | | 0.66 (0.40-1.08) | | 0.48 (0.27-0.85) | |  |
| **Winter** | | 105/11109 (0.9%) | | 0.94 (0.58-1.51) | | 0.69 (0.39-1.21) | | 0.42 (0.20-0.90) | |  |
| **Hypertension** | |  | |  | |  | |  | | 0.991 |
| **No** | | 206/27140 (0.8%) | | 0.71 (0.50-1.02) | | 0.63 (0.42-0.92) | | 0.43 (0.27-0.69) | |  |
| **Yes** | | 266/24577 (1.1%) | | 0.64 (0.47-0.88) | | 0.61 (0.43-0.86) | | 0.38 (0.25-0.60) | |  |

Multivariable Cox proportional hazard models were used.Model was adjusted for age, sex, ethnicity, townsend deprivation index, assessment centre, education, sleep duration, smoking status,alcohol consumption, BMI, season of accelerometer wearing, hypertension, platelet count, ALT, AST, cholesterol, and triglycerides.

**Table S5. HR (95%CI) for incident MASLD associated with plasma n-6 PUFA in various subgroups.**

| **Subgroups** | **Events/n(%)** | | **n-6 PUFA Q2 (36.2-38.7)** | | **n-6 PUFA Q3 (38.7-40.7)** | | **n-6 PUFA Q4 (40.7-50.3)** | | ***P* for interaction** | |
| --- | --- | --- | --- | --- | --- | --- | --- | --- | --- | --- |
| **Sex** |  | |  | |  | |  | | 0.062 | |
| **Female** | 248/28882 (0.9%) | | 0.89 (0.62-1.28) | | 0.78 (0.51-1.19) | | 0.38 (0.22-0.67) | |  | |
| **Male** | 224/22835 (1%) | | 0.52 (0.35-0.76) | | 0.50 (0.31-0.79) | | 0.52 (0.31-0.87) | |  | |
| **Age,years** |  | |  | |  | |  | | 0.502 | |
| **< 60** | 295/30838 (1%) | | 0.75 (0.54-1.05) | | 0.68 (0.46-0.99) | | 0.42 (0.26-0.66) | |  | |
| **≥ 60** | 177/20879 (0.8%) | | 0.59 (0.39-0.89) | | 0.54 (0.33-0.88) | | 0.47 (0.26-0.86) | |  | |
| **Ethnicity** |  | |  | |  | |  | | 0.325 | |
| **Nonwhite** | 22/1454 (1.5%) | | 3.42 (0.66-17.64) | | 9.17 (1.84-45.76) | | 5.95 (0.92-38.37) | |  | |
| **White** | 450/50263 (0.9%) | | 0.65 (0.50-0.84) | | 0.56 (0.41-0.76) | | 0.38 (0.26-0.57) | |  | |
| **Recruitment center** |  | |  | |  | |  | | 0.423 | |
| **England** | 452/47461 (1%) | | 0.69 (0.53-0.89) | | 0.64 (0.47-0.86) | | 0.40 (0.27-0.59) | |  | |
| **Scotland** | 10/2748 (0.4%) | | 0.98 (0.11-8.45) | | 1.04 (0.10-11.18) | | 1.71 (0.12-24.37) | |  | |
| **Wales** | 10/1508 (0.7%) | | 0.60 (0.09-3.82) | | 0.00 (0.00-0.00) | | 1.68 (0.17-16.23) | |  | |
| **education** |  | |  | |  | |  | | 0.878 | |
| **Any other qualification** | 266/25105 (1.1%) | | 0.69 (0.49-0.98) | | 0.64 (0.43-0.95) | | 0.52 (0.32-0.84) | |  | |
| **Degree or above** | 150/22030 (0.7%) | | 0.69 (0.44-1.08) | | 0.56 (0.33-0.95) | | 0.33 (0.17-0.65) | |  | |
| **No qualification** | 56/4582 (1.2%) | | 0.67 (0.32-1.42) | | 0.76 (0.32-1.79) | | 0.21 (0.05-0.84) | |  | |
| **Smoking status** |  | |  | |  | |  | | 0.545 | |
| **Current** | 55/3538 (1.6%) | | 1.16 (0.53-2.55) | | 1.50 (0.62-3.68) | | 1.10 (0.35-3.43) | |  | |
| **Never** | 217/29528 (0.7%) | | 0.76 (0.52-1.13) | | 0.68 (0.43-1.06) | | 0.54 (0.32-0.91) | |  | |
| **Previous** | 200/18651 (1.1%) | | 0.53 (0.36-0.78) | | 0.47 (0.29-0.76) | | 0.25 (0.13-0.47) | |  | |
| **Alcohol consumption** |  | |  | |  | |  | | 0.789 | |
| **<3 times/week** | 238/23539 (1%) | | 0.60 (0.41-0.87) | | 0.59 (0.38-0.90) | | 0.42 (0.26-0.70) | |  | |
| **3+ times/week** | | 186/25297 (0.7%) | | 0.68 (0.45-1.01) | | 0.55 (0.34-0.89) | | 0.34 (0.18-0.65) | |  |
| **Never** | | 48/2881 (1.7%) | | 1.44 (0.61-3.40) | | 1.55 (0.59-4.10) | | 0.99 (0.31-3.16) | |  |
| **BMI,kg/m^2^** | |  | |  | |  | |  | | 0.245 |
| **< 25** | | 61/19957 (0.3%) | | 0.53 (0.24-1.18) | | 0.56 (0.25-1.29) | | 0.27 (0.10-0.72) | |  |
| **≥ 25** | | 411/31760 (1.3%) | | 0.70 (0.53-0.92) | | 0.64 (0.46-0.88) | | 0.52 (0.35-0.78) | |  |
| **Season of accelerometer wearing** | |  | |  | |  | |  | | 0.947 |
| **Autumn** | | 153/15784 (1%) | | 0.59 (0.37-0.95) | | 0.69 (0.41-1.16) | | 0.41 (0.22-0.78) | |  |
| **Spring** | | 83/11354 (0.7%) | | 0.82 (0.45-1.51) | | 0.63 (0.30-1.32) | | 0.56 (0.24-1.33) | |  |
| **Summer** | | 131/13470 (1%) | | 0.67 (0.41-1.10) | | 0.62 (0.35-1.10) | | 0.50 (0.25-0.99) | |  |
| **Winter** | | 105/11109 (0.9%) | | 0.77 (0.46-1.30) | | 0.57 (0.30-1.09) | | 0.37 (0.16-0.85) | |  |
| **Hypertension** | |  | |  | |  | |  | | 0.091 |
| **No** | | 206/27140 (0.8%) | | 0.56 (0.37-0.86) | | 0.75 (0.48-1.17) | | 0.54 (0.32-0.91) | |  |
| **Yes** | | 266/24577 (1.1%) | | 0.78 (0.56-1.08) | | 0.56 (0.37-0.85) | | 0.35 (0.20-0.60) | |  |

Multivariable Cox proportional hazard models were used.Model was adjusted for age, sex, ethnicity, townsend deprivation index, assessment centre, education, sleep duration, smoking status,alcohol consumption, BMI, season of accelerometer wearing, hypertension, platelet count, ALT, AST, cholesterol, and triglycerides.

**Table S6. HR (95%CI) for incident MASLD associated with plasma MUFA in various subgroups.**

| **Subgroups** | **Events/n(%)** | **MUFA Q2 (21.6-23.1)** | **MUFA Q3 (23.1-25.0)** | **MUFA Q4 (25.0-37.9)** | ***P* for interaction** |
| --- | --- | --- | --- | --- | --- |
| **Sex** |  |  |  |  | 0.101 |
| **Female** | 248/28882 (0.9%) | 1.52 (0.99-2.36) | 1.61 (1.04-2.49) | 1.50 (0.87-2.56) |  |
| **Male** | 224/22835 (1%) | 0.64 (0.36-1.17) | 1.02 (0.61-1.68) | 1.32 (0.78-2.23) |  |
| **Age,years** |  |  |  |  | 0.215 |
| **< 60** | 295/30838 (1%) | 1.30 (0.82-2.07) | 1.68 (1.09-2.61) | 1.81 (1.11-2.95) |  |
| **≥ 60** | 177/20879 (0.8%) | 0.99 (0.59-1.68) | 1.08 (0.65-1.80) | 1.33 (0.74-2.38) |  |
| **Ethnicity** |  |  |  |  | 0.195 |
| **Nonwhite** | 22/1454 (1.5%) | 1.49 (0.47-4.76) | 0.31 (0.07-1.39) | 0.18 (0.03-0.98) |  |
| **White** | 450/50263 (0.9%) | 1.14 (0.79-1.65) | 1.51 (1.07-2.14) | 1.75 (1.19-2.58) |  |
| **Recruitment center** |  |  |  |  | 0.130 |
| **England** | 452/47461 (1%) | 1.10 (0.77-1.57) | 1.40 (1.00-1.95) | 1.59 (1.09-2.32) |  |
| **Scotland** | 10/2748 (0.4%) | 0.00 (0.00-0.00) | 0.00 (0.00-0.00) | 0.00 (0.00-0.00) |  |
| **Wales** | 10/1508 (0.7%) | 0.45 (0.02-8.35) | 1.19 (0.11-12.43) | 0.28 (0.02-4.81) |  |
| **education** |  |  |  |  | 0.576 |
| **Any other qualification** | 266/25105 (1.1%) | 1.50 (0.90-2.50) | 1.77 (1.09-2.87) | 2.23 (1.32-3.77) |  |
| **Degree or above** | 150/22030 (0.7%) | 0.90 (0.52-1.56) | 1.13 (0.67-1.93) | 1.29 (0.68-2.43) |  |
| **No qualification** | 56/4582 (1.2%) | 0.89 (0.31-2.57) | 1.11 (0.42-2.96) | 0.69 (0.22-2.13) |  |
| **Smoking status** |  |  |  |  | 0.322 |
| **Current** | 55/3538 (1.6%) | 0.54 (0.17-1.74) | 0.59 (0.21-1.67) | 0.59 (0.20-1.73) |  |
| **Never** | 217/29528 (0.7%) | 1.42 (0.89-2.29) | 1.50 (0.94-2.41) | 1.46 (0.84-2.55) |  |
| **Previous** | 200/18651 (1.1%) | 0.89 (0.50-1.58) | 1.50 (0.89-2.52) | 2.08 (1.18-3.67) |  |
| **Alcohol consumption** |  |  |  |  | 0.664 |
| **<3 times/week** | 238/23539 (1%) | 0.92 (0.55-1.56) | 1.28 (0.78-2.08) | 1.75 (1.03-2.98) |  |

| **3+ times/week** | 186/25297 (0.7%) | 1.17 (0.70-1.96) | 1.50 (0.92-2.45) | 1.45 (0.83-2.54) |  |
| --- | --- | --- | --- | --- | --- |
| **Never** | 48/2881 (1.7%) | 1.97 (0.62-6.29) | 1.38 (0.43-4.49) | 1.05 (0.27-4.05) |  |
| **BMI,kg/m^2^** |  |  |  |  | 0.318 |
| **< 25** | 61/19957 (0.3%) | 1.55 (0.76-3.18) | 1.82 (0.83-3.97) | 2.37 (0.82-6.85) |  |
| **≥ 25** | 411/31760 (1.3%) | 0.94 (0.63-1.40) | 1.12 (0.78-1.62) | 1.26 (0.84-1.88) |  |
| **Season of accelerometer wearing** |  |  |  |  | 0.614 |
| **Autumn** | 153/15784 (1%) | 1.32 (0.74-2.35) | 1.30 (0.73-2.31) | 1.46 (0.76-2.82) |  |
| **Spring** | 83/11354 (0.7%) | 1.34 (0.62-2.89) | 1.14 (0.53-2.47) | 0.97 (0.40-2.35) |  |
| **Summer** | 131/13470 (1%) | 1.07 (0.53-2.16) | 1.47 (0.77-2.80) | 1.72 (0.84-3.52) |  |
| **Winter** | 105/11109 (0.9%) | 0.70 (0.31-1.58) | 1.46 (0.73-2.94) | 2.01 (0.92-4.40) |  |
| **Hypertension** |  |  |  |  | 0.365 |
| **No** | 206/27140 (0.8%) | 0.96 (0.58-1.59) | 1.29 (0.81-2.06) | 1.30 (0.75-2.24) |  |
| **Yes** | 266/24577 (1.1%) | 1.33 (0.82-2.18) | 1.49 (0.93-2.38) | 1.76 (1.05-2.95) |  |

Multivariable Cox proportional hazard models were used.Model was adjusted for age, sex, ethnicity, townsend deprivation index, assessment centre, education, sleep duration, smoking status,alcohol consumption, BMI, season of accelerometer wearing, hypertension, platelet count, ALT, AST, cholesterol, and triglycerides.

**Table S7. HR (95%CI) for incident MASLD associated with plasma SFA in various subgroups.**

| **Subgroups** | **Events/n(%)** | **SFA Q2**  **(32.7-33.8)** | **SFA Q3**  **(33.8-35.0)** | **SFA Q4**  **(35.0-45.5)** | ***P* for interaction** |
| --- | --- | --- | --- | --- | --- |
| **Sex** |  |  |  |  | 0.653 |
| **Female** | 248/28882 (0.9%) | 1.64 (1.09-2.47) | 1.50 (0.99-2.27) | 1.81 (1.19-2.75) |  |
| **Male** | 224/22835 (1%) | 1.18 (0.74-1.88) | 1.44 (0.92-2.25) | 1.94 (1.28-2.94) |  |
| **Age,years** |  |  |  |  | 0.466 |
| **< 60** | 295/30838 (1%) | 1.25 (0.85-1.83) | 1.28 (0.88-1.88) | 1.82 (1.27-2.62) |  |
| **≥ 60** | 177/20879 (0.8%) | 1.76 (1.05-2.95) | 1.86 (1.12-3.10) | 2.09 (1.27-3.45) |  |
| **Ethnicity** |  |  |  |  | 0.962 |
| **Nonwhite** | 22/1454 (1.5%) | 1.30 (0.39-4.29) | 1.24 (0.31-5.02) | 0.57 (0.12-2.60) |  |
| **White** | 450/50263 (0.9%) | 1.44 (1.05-1.98) | 1.49 (1.08-2.03) | 1.96 (1.45-2.66) |  |
| **Recruitment center** |  |  |  |  | 0.990 |
| **England** | 452/47461 (1%) | 1.41 (1.04-1.93) | 1.45 (1.06-1.97) | 1.85 (1.37-2.49) |  |
| **Scotland** | 10/2748 (0.4%) | 1.43 (0.12-16.55) | 1.59 (0.13-18.86) | 2.71 (0.26-28.68) |  |
| **Wales** | 10/1508 (0.7%) | 2.53 (0.17-37.84) | 3.09 (0.20-47.52) | 6.45 (0.45-92.60) |  |
| **education** |  |  |  |  | 0.056 |
| **Any other qualification** | 266/25105 (1.1%) | 1.49 (1.01-2.21) | 1.14 (0.76-1.72) | 1.64 (1.11-2.41) |  |
| **Degree or above** | 150/22030 (0.7%) | 1.17 (0.69-2.00) | 1.50 (0.90-2.49) | 1.85 (1.12-3.05) |  |
| **No qualification** | 56/4582 (1.2%) | 2.42 (0.59-9.84) | 7.12 (2.03-24.94) | 7.17 (2.05-25.11) |  |
| **Smoking status** |  |  |  |  | 0.038 |
| **Current** | 55/3538 (1.6%) | 2.75 (0.97-7.75) | 2.35 (0.84-6.58) | 1.55 (0.53-4.49) |  |
| **Never** | 217/29528 (0.7%) | 1.24 (0.83-1.85) | 1.06 (0.69-1.61) | 1.47 (0.98-2.21) |  |
| **Previous** | 200/18651 (1.1%) | 1.50 (0.86-2.60) | 1.94 (1.16-3.25) | 2.96 (1.81-4.83) |  |
| **Alcohol consumption** |  |  |  |  | 0.444 |
| **<3 times/week** | 238/23539 (1%) | 1.39 (0.94-2.06) | 1.19 (0.79-1.80) | 1.65 (1.11-2.46) |  |
| **3+ times/week** | 186/25297 (0.7%) | 1.56 (0.84-2.90) | 2.24 (1.26-3.95) | 2.77 (1.59-4.81) |  |
| **Never** | 48/2881 (1.7%) | 1.31 (0.57-2.99) | 1.10 (0.46-2.64) | 0.96 (0.39-2.32) |  |
| **BMI,kg/m^2^** |  |  |  |  | 0.13 |
| **< 25** | 61/19957 (0.3%) | 1.74 (0.80-3.80) | 1.21 (0.50-2.90) | 3.20 (1.49-6.89) |  |
| **≥ 25** | 411/31760 (1.3%) | 1.36 (0.98-1.90) | 1.46 (1.06-2.03) | 1.77 (1.29-2.43) |  |
| **Season of accelerometer wearing** |  |  |  |  | 0.844 |
| **Autumn** | 153/15784 (1%) | 1.68 (0.99-2.84) | 1.34 (0.77-2.33) | 2.03 (1.20-3.42) |  |
| **Spring** | 83/11354 (0.7%) | 1.45 (0.72-2.92) | 1.52 (0.76-3.06) | 1.57 (0.79-3.12) |  |
| **Summer** | 131/13470 (1%) | 1.24 (0.68-2.25) | 1.28 (0.71-2.31) | 2.00 (1.16-3.47) |  |
| **Winter** | 105/11109 (0.9%) | 1.21 (0.61-2.39) | 1.69 (0.91-3.17) | 1.88 (1.00-3.54) |  |
| **Hypertension** |  |  |  |  | 0.069 |
| **No** | 206/27140 (0.8%) | 1.00 (0.66-1.53) | 1.06 (0.70-1.62) | 1.52 (1.02-2.28) |  |
| **Yes** | 266/24577 (1.1%) | 2.16 (1.35-3.44) | 2.15 (1.35-3.42) | 2.58 (1.64-4.04) |  |

Multivariable Cox proportional hazard models were used.Model was adjusted for age, sex, ethnicity, townsend deprivation index, assessment centre, education, sleep duration, smoking status,alcohol consumption, BMI, season of accelerometer wearing, hypertension, platelet count, ALT, AST, cholesterol, and triglycerides.

**Table S8. Sensitivity analysis of the associations between MVPA，plasma n-6 PUFA，MUFA，SFA levels and incident MASLD risk by excluding participants within 2 years of follow-up.**

| **Exposure Group** | **Events/n(rate^a^)** | **Model 1** |  | **Model 2** |  | **Model 3** |  |
| --- | --- | --- | --- | --- | --- | --- | --- |
|  |  | **HR (95% CI)** | ***P* value** | **HR (95% CI)** | ***P* value** | **HR (95% CI)** | ***P* value** |
| **MVPA(min/week)** |  |  |  |  |  |  |  |
| **Q1** | 182/12817 (1.9) | 1(Ref) |  | 1(Ref) |  | 1(Ref) |  |
| **Q2** | 92/12807 (0.9) | 0.47 (0.36-0.60) | <0.001 | 0.62 (0.48-0.81) | <0.001 | 0.65 (0.50-0.84) | <0.001 |
| **Q3** | 77/12849 (0.8) | 0.38 (0.29-0.49) | <0.001 | 0.56 (0.43-0.75) | <0.001 | 0.61 (0.46-0.80) | <0.001 |
| **Q4** | 44/12885 (0.4) | 0.21 (0.15-0.29) | <0.001 | 0.35 (0.24-0.49) | <0.001 | 0.38 (0.27-0.54) | <0.001 |
| **n-6 PUFA(%TFA)** |  |  |  |  |  |  |  |
| **Q1** | 185/13047 (1.8) | 1(Ref) |  | 1(Ref) |  | 1(Ref) |  |
| **Q2** | 94/12838 (0.9) | 0.50 (0.39-0.65) | <0.001 | 0.63 (0.49-0.81) | <0.001 | 0.74 (0.56-0.98) | 0.034 |
| **Q3** | 70/13019 (0.7) | 0.36 (0.27-0.48) | <0.001 | 0.53 (0.40-0.70) | <0.001 | 0.66 (0.47-0.91) | 0.012 |
| **Q4** | 46/12454 (0.5) | 0.23 (0.17-0.32) | <0.001 | 0.39 (0.28-0.55) | <0.001 | 0.48 (0.33-0.72) | <0.001 |
| **MUFA(%TFA)** |  |  |  |  |  |  |  |
| **Q1** | 43/12863 (0.4) | 1(Ref) |  | 1(Ref) |  | 1(Ref) |  |
| **Q2** | 73/13217 (0.7) | 1.70 (1.17-2.48) | 0.006 | 1.39 (0.95-2.03) | 0.089 | 1.35 (0.92-1.98) | 0.125 |
| **Q3** | 106/12935 (1.0) | 2.60 (1.82-3.71) | <0.001 | 1.69 (1.18-2.44) | 0.005 | 1.52 (1.04-2.21) | 0.030 |
| **Q4** | 173/12343 (1.8) | 4.59 (3.26-6.46) | <0.001 | 2.32 (1.62-3.33) | <0.001 | 1.69 (1.11-2.57) | 0.014 |
| **SFA(%TFA)** |  |  |  |  |  |  |  |
| **Q1** | 67/13406 (0.6) | 1(Ref) |  | 1(Ref) |  | 1(Ref) |  |
| **Q2** | 79/12766 (0.8) | 1.27 (0.92-1.76) | 0.148 | 1.24 (0.89-1.72) | 0.199 | 1.24 (0.90-1.73) | 0.192 |
| **Q3** | 90/12520 (0.9) | 1.49 (1.08-2.04) | 0.014 | 1.39 (1.01-1.91) | 0.045 | 1.31 (0.95-1.81) | 0.099 |
| **Q4** | 159/12666 (1.6) | 2.61 (1.96-3.48) | <0.001 | 2.07 (1.54-2.78) | <0.001 | 1.68 (1.23-2.30) | 0.001 |

^a^Incidence rates were expressed as per 1000 person-years

Multivariable Cox proportional hazard models were used:Model 1 was adjusted for age, sex, and ethnicity.Model 2 was adjusted for Model 1 plus Townsend deprivation index, assessment centre, education, sleep duration, smoking status,alcohol consumption, BMI, and season of accelerometer wearing.Model 3 was adjusted for Model 2 plus hypertension, platelet count, ALT, AST, cholesterol, and triglycerides.

**Table S9. Sensitivity analysis of the associations between MVPA，plasma n-6 PUFA，MUFA，SFA levels and incident MASLD risk using a complete-case dataset without missing values.**

| **Exposure Group** | **Events/n(rate^a^)** | **Model 1** |  | **Model 2** |  | **Model 3** |  |
| --- | --- | --- | --- | --- | --- | --- | --- |
|  |  | **HR (95% CI)** | ***P* value** | **HR (95% CI)** | ***P* value** | **HR (95% CI)** | ***P* value** |
| **MVPA(min/week)** |  |  |  |  |  |  |  |
| **Q1** | 182/11344 (2.1) | 1(Ref) |  | 1(Ref) |  | 1(Ref) |  |
| **Q2** | 100/11289 (1.1) | 0.51 (0.40-0.65) | <0.001 | 0.67 (0.52-0.86) | 0.002 | 0.71 (0.55-0.91) | 0.007 |
| **Q3** | 78/11314 (0.9) | 0.38 (0.29-0.50) | <0.001 | 0.57 (0.43-0.75) | <0.001 | 0.62 (0.47-0.82) | <0.001 |
| **Q4** | 46/11235 (0.5) | 0.22 (0.16-0.30) | <0.001 | 0.36 (0.25-0.51) | <0.001 | 0.40 (0.28-0.57) | <0.001 |
| **n-6 PUFA(%TFA)** |  |  |  |  |  |  |  |
| **Q1** | 196/11510 (2.2) | 1(Ref) |  | 1(Ref) |  | 1(Ref) |  |
| **Q2** | 94/11320 (1.1) | 0.48 (0.37-0.61) | <0.001 | 0.59 (0.46-0.76) | <0.001 | 0.69 (0.52-0.92) | 0.011 |
| **Q3** | 73/11436 (0.8) | 0.36 (0.27-0.47) | <0.001 | 0.52 (0.39-0.69) | <0.001 | 0.65 (0.46-0.90) | 0.009 |
| **Q4** | 43/10916 (0.5) | 0.21 (0.15-0.29) | <0.001 | 0.34 (0.24-0.48) | <0.001 | 0.43 (0.28-0.64) | <0.001 |
| **MUFA(%TFA)** |  |  |  |  |  |  |  |
| **Q1** | 53/11978 (0.6) | 1(Ref) |  | 1(Ref) |  | 1(Ref) |  |
| **Q2** | 60/10924 (0.7) | 1.27 (0.88-1.84) | 0.201 | 1.05 (0.72-1.52) | 0.817 | 0.99 (0.68-1.44) | 0.948 |
| **Q3** | 109/11425 (1.2) | 2.27 (1.63-3.16) | <0.001 | 1.50 (1.07-2.10) | 0.020 | 1.30 (0.91-1.84) | 0.147 |
| **Q4** | 184/10855 (2.2) | 4.15 (3.03-5.68) | <0.001 | 2.13 (1.52-2.97) | <0.001 | 1.49 (1.00-2.23) | 0.051 |
| **SFA(%TFA)** |  |  |  |  |  |  |  |
| **Q1** | 65/11829 (0.7) | 1(Ref) |  | 1(Ref) |  | 1(Ref) |  |
| **Q2** | 81/11159 (0.9) | 1.36 (0.98-1.89) | 0.064 | 1.31 (0.94-1.81) | 0.111 | 1.30 (0.94-1.81) | 0.119 |
| **Q3** | 90/11049 (1.1) | 1.54 (1.12-2.12) | 0.008 | 1.42 (1.03-1.96) | 0.034 | 1.33 (0.96-1.84) | 0.087 |
| **Q4** | 170/11145 (2.0) | 2.89 (2.17-3.86) | <0.001 | 2.28 (1.70-3.07) | <0.001 | 1.82 (1.33-2.49) | <0.001 |

^a^Incidence rates were expressed as per 1000 person-years

Multivariable Cox proportional hazard models were used:Model 1 was adjusted for age, sex, and ethnicity.Model 2 was adjusted for Model 1 plus Townsend

deprivation index, assessment centre, education, sleep duration, smoking status,alcohol consumption, BMI, and season of accelerometer wearing.Model 3 was adjusted for Model 2 plus hypertension, platelet count, ALT, AST, cholesterol, and triglycerides.

**Table S10. Sensitivity analysis of the associations between MVPA，plasma n-6 PUFA，MUFA，SFA levels and incident MASLD risk by excluding participants with hypertension or diabetes at baseline.**

| **Exposure Group** | **Events/n(rate^a^)** | **Model 1** |  | **Model 2** |  | **Model 3** |  |
| --- | --- | --- | --- | --- | --- | --- | --- |
|  |  | **HR (95% CI)** | ***P* value** | **HR (95% CI)** | ***P* value** | **HR (95% CI)** | ***P* value** |
| **MVPA(min/week)** |  |  |  |  |  |  |  |
| **Q1** | 73/5661 (1.7) | 1(Ref) |  | 1(Ref) |  | 1(Ref) |  |
| **Q2** | 48/6460 (0.7) | 0.53 (0.37-0.77) | <0.001 | 0.70 (0.48-1.02) | 0.062 | 0.70 (0.48-1.02) | 0.066 |
| **Q3** | 44/7041 (0.6) | 0.43 (0.29-0.63) | <0.001 | 0.64 (0.43-0.95) | 0.027 | 0.64 (0.43-0.95) | 0.028 |
| **Q4** | 27/7500 (0.4) | 0.24 (0.15-0.37) | <0.001 | 0.42 (0.26-0.67) | <0.001 | 0.45 (0.28-0.72) | 0.008 |
| **n-6 PUFA(%TFA)** |  |  |  |  |  |  |  |
| **Q1** | 83/5035 (2.1) | 1(Ref) |  | 1(Ref) |  | 1(Ref) |  |
| **Q2** | 36/6229 (0.7) | 0.34 (0.23-0.51) | <0.001 | 0.43 (0.29-0.64) | <0.001 | 0.54 (0.35-0.84) | 0.007 |
| **Q3** | 41/7293 (0.7) | 0.33 (0.22-0.48) | <0.001 | 0.51 (0.34-0.75) | 0.001 | 0.70 (0.44-1.11) | 0.130 |
| **Q4** | 32/8105 (0.5) | 0.22 (0.14-0.33) | <0.001 | 0.38 (0.25-0.59) | <0.001 | 0.54 (0.32-0.91) | 0.021 |
| **MUFA(%TFA)** |  |  |  |  |  |  |  |
| **Q1** | 31/8522 (0.5) | 1(Ref) |  | 1(Ref) |  | 1(Ref) |  |
| **Q2** | 32/6782 (0.6) | 1.32 (0.81-2.17) | 0.266 | 1.05 (0.64-1.73) | 0.843 | 0.97 (0.58-1.60) | 0.894 |
| **Q3** | 52/6290 (1.1) | 2.38 (1.52-3.72) | <0.001 | 1.47 (0.93-2.34) | 0.101 | 1.21 (0.75-1.95) | 0.442 |
| **Q4** | 77/5068 (2.0) | 4.47 (2.91-6.88) | <0.001 | 2.05 (1.29-3.25) | 0.002 | 1.22 (0.70-2.13) | 0.492 |
| **SFA(%TFA)** |  |  |  |  |  |  |  |
| **Q1** | 44/7947 (0.7) | 1(Ref) |  | 1(Ref) |  | 1(Ref) |  |
| **Q2** | 39/7095 (0.7) | 1.00 (0.65-1.55) | 0.983 | 1.00 (0.65-1.54) | 0.999 | 1.03 (0.66-1.59) | 0.906 |
| **Q3** | 42/6378 (0.8) | 1.21 (0.79-1.84) | 0.386 | 1.16 (0.75-1.77) | 0.507 | 1.10 (0.71-1.69) | 0.675 |
| **Q4** | 67/5242 (1.7) | 2.35 (1.60-3.44) | <0.001 | 1.95 (1.32-2.88) | <0.001 | 1.55 (1.02-2.36) | <0.001 |

^a^Incidence rates were expressed as per 1000 person-years

Multivariable Cox proportional hazard models were used:Model 1 was adjusted for age, sex, and ethnicity.Model 2 was adjusted for Model 1 plus Townsend

deprivation index, assessment centre, education, sleep duration, smoking status,alcohol consumption, BMI, and season of accelerometer wearing.Model 3 was adjusted for Model 2 plus platelet count, ALT, AST, cholesterol, and triglycerides.

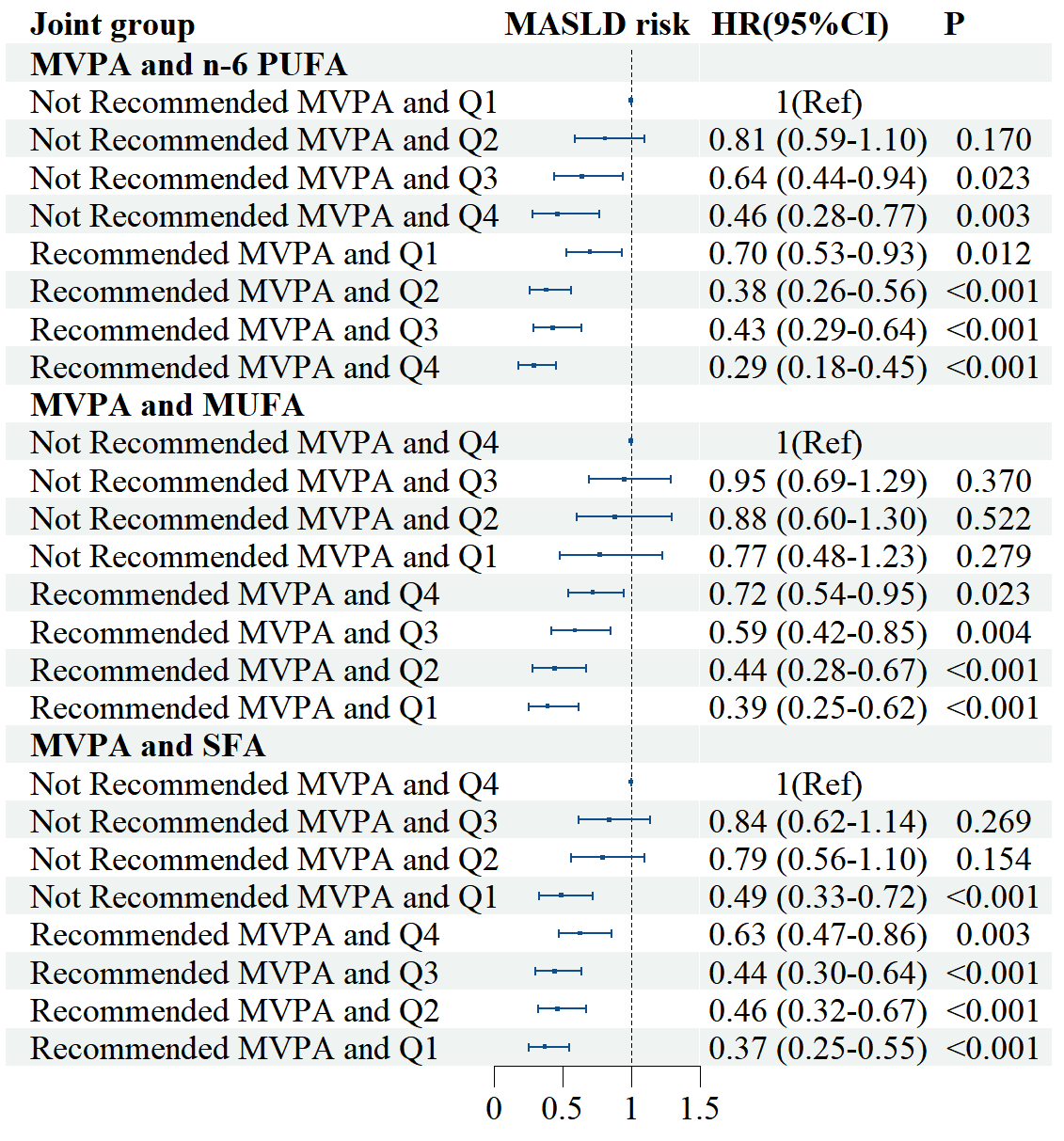

**Fig S1****.The joint association of MVPA and fatty acids with the risk of MASLD.**

According to the WHO guidelines, MVPA was categorized into two groups: Recommended MVPA (MVPA ≥ 150 minutes/week) and Not Recommended MVPA (MVPA < 150 minutes/week). The multivariable Cox model was adjusted for age, sex, ethnicity, Townsend deprivation index, assessment center, education, sleep duration, smoking status, alcohol consumption, BMI, season of accelerometer wearing, hypertension, platelet count, ALT, AST, cholesterol, and triglycerides. 95% CI = 95% confidence interval; MVPA = moderate to vigorous physical activity; WHO = World Health Organization.
